# Computational framework for the World Health Organization estimates of the global, regional and national burden of foodborne diseases 2026 edition

**DOI:** 10.64898/2026.05.13.26353030

**Authors:** Brecht Devleesschauwer, Louise Vaes, Kim Fernandez, Elaine Borghi, Bochen Cao, Christina Fastl, Lea Jakobsen, Richard Kumapley, Rob Lake, Shannon E Majowicz, Yuki Minato, Sara Monteiro Pires, Lapo Mughini-Gras, Gabriela F Nane, Lucy J Robertson, Elaine Scallan Walter, Paul R Torgerson, Mirjam Kretzschmar, Carlotta di Bari

## Abstract

**Background:** Foodborne diseases cause substantial global morbidity and mortality, yet remain largely unattended. To support countries to address this public health concern, the World Health Assembly Resolution 73.5 called for strengthening global food safety efforts and led to the development of the WHO Global Strategy for Food Safety 2022–2030, adopted at the 75th WHA (2022). To this end, the World Health Organization (WHO) reconvened the Foodborne Disease Burden Epidemiology Reference Group (FERG) to advise and support the work to generate updated global, regional, and national estimates of the foodborne disease burden for the reference period 2000–2021.

**Methods:** We developed an incidence-based framework expanding coverage to 42 foodborne hazards. Standardized systematic reviews, Global Health Estimates and Global Burden of Disease envelopes, and United Nations population data informed the evidence base. Missing epidemiological data were imputed using Bayesian hierarchical meta-regression models. Disease models mapped acute and chronic health outcomes, applying updated disability weights, life tables, and probabilistic Monte Carlo calculations to estimate incidence, mortality, Years Lived with Disability, Years of Life Lost and Disability-Adjusted Life Years for all 194 WHO Member States. Transparency and analysis reproducibility were ensured through availed open source R packages and standardized workflows.

**Results:** The computational framework provides annual, country-level estimates with improved internal consistency and an expanded hazard scope compared with the WHO 2015 edition. Advances include refined modelling, enhanced uncertainty propagation, and broader inclusion of microbial, parasitic, and chemical hazards. Persistent data gaps—especially in high-burden regions—were filled through extensive imputation.

**Conclusions:** The computational framework for the WHO 2026 edition delivers the most comprehensive and transparent assessment of the global burden of foodborne diseases to date. Despite remaining limitations, it enables routine monitoring, supports evaluation of global food safety efforts, and highlights priorities for strengthening national data systems.

## Introduction

In 2015, the World Health Organization (WHO), with technical support from the Foodborne Disease Burden Epidemiology Reference Group (FERG), published the first-ever estimates of the global burden of foodborne diseases, documenting the population health impact of 31 foodborne hazards in the year 2010 [1]. These estimates uncovered the hidden burden of unsafe food, with an estimated number of 600 million illnesses, 420 thousand deaths, and 33 million disability-adjusted life years (DALYs) worldwide. Vulnerable populations were found to be disproportionally affected by foodborne diseases, with children <5 years of age accounting for one third of all foodborne deaths. These landmark estimates placed food safety on the global agenda, and sparked interest in assessing the economic burden of foodborne diseases [2] and national initiatives on fine tuning estimates and surveillance systems [3].

In 2020, the WHO Member States mandated the WHO via the Seventy-third World Health Assembly resolution on “Strengthening efforts on food safety” (WHA73.5), to regularly monitor and report on the global burden of foodborne and zoonotic diseases at national, regional and international levels. They also mandated the preparation of a new report by 2025 on the global burden of foodborne diseases with up-to-date estimates of global foodborne disease incidence, mortality and disease burden in terms of DALYs. This revamped monitoring would also allow for measuring the impact of the WHO Global Strategy for Food Safety 2022–2030. To achieve these objectives, WHO reconvened FERG in 2021 for the 2021–2025 period. The group comprised 26 members organised into seven task forces (TFs) based on their main areas of expertise. These included the Enteric Diseases TF (EDTF), the Parasitic Diseases TF (PDTF), and the Chemicals and Toxins TF (CTTF). The Source Attribution TF (SATF) was responsible for assessing the relative contributions of different transmission routes and foods to the foodborne disease burden. The Country Support TF (CSTF) and the Impact Measurement TF (IMTF) addressed the expanded mandate of FERG. Finally, the Computational TF (CTF) oversaw the establishment and implementation of the computational framework underlying the development of the updated estimates. WHO commissioned Sciensano, the Belgian institute for health, for carrying out the computational work, under WHO’s supervision and advice from the CTF.

The computational framework used here builds on the approach developed for the initial estimates of the global burden of foodborne diseases circa 2010 [4], but comes with improvements in both methods and application. As before, the hazard- and incidence-based approach was adopted for defining the burden of foodborne diseases [5]. Compared with the previous iteration, the current framework allowed for generating country and year-specific estimates, and improved internal consistency and transparency.

This manuscript documents the overall computational framework of the WHO 2026 edition estimates of the global, regional and national burden of foodborne disease (Figure 1). The description of the framework adheres to the STROBOD and GATHER statements for reporting on burden of disease studies and global health estimates, respectively [6–7] (**S1 Appendix**, **S2 Appendix**). Additional details on the hazard-specific methods, as well as on the approaches for source attribution, are documented elsewhere [8–13].

**Figure 1.**
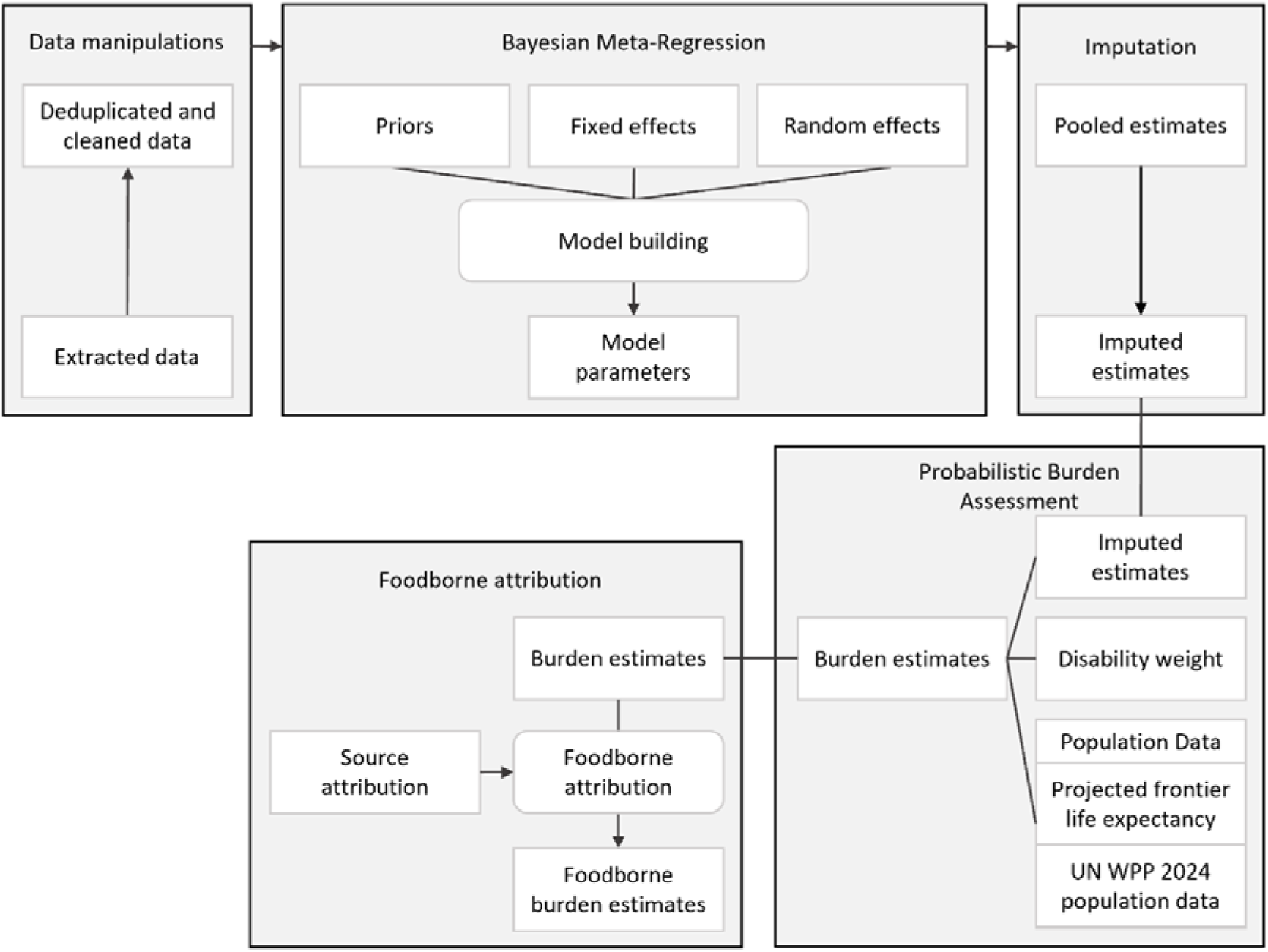
Computational framework for World Health Organization estimates of the global burden of foodborne diseases 2026 edition

## Study setting

### Hazards

The WHO 2015 edition estimates covered the global burden of 31 foodborne hazards. The burden of six additional hazards could only be estimated in high-income regions. With the updated WHO 2026 edition estimates, we aimed to increase the scope by expanding the number of hazards in function of the increased availability of data and resources.

To support the hazard selection activity, a hazard prioritization matrix was developed (**S3 Appendix**). This tool did not serve as a mechanistic decision algorithm, but rather aimed to document the implicit choices that are made when prioritizing hazards for inclusion in the estimation framework. The hazard-specific TFs were able to enhance the prioritization matrix to better suit their specificities by adding criteria specific to the TF.

The tool comprised of a minimum set of six dimensions, scored qualitatively on an ordinal scale, based on current evidence (**S3 Appendix**). When data were not available to support the evaluation, scores relied on expert opinion. The term “estimated” in the dimension definitions therefore refers to both data-driven and expert-driven assessments. In addition, expert judgement was sought to evaluate dimensions as high, medium or low, with indicative definitions. However, full quantification of these scores was not possible due to variation across hazards. Finally, while the tool did not required dimensions to be combined into an overall score, some hazard-specific TFs did so as part of their decision-making process. Individual TF scoring and results are documented in separate papers [9–12].

In the end, 42 hazards were selected, including 14 diarrhoeal pathogens, 7 invasive enteric, 12 invasive parasitic and 9 chemical hazards [14]. Compared to the WHO 2015 edition estimates, novel hazards included *Trypanosoma cruzi* (agent of Chagas disease), *Cyclospora* spp., rotavirus, enteroaggregative *Escherichia coli* (EAEC), aflatoxin M1, and four foodborne metals (arsenic, cadmium, methylmercury, and lead), for which estimates were previously published in a standalone effort [15]. *Clostridium perfringens*, *Bacillus cereus*, and *Staphylococcus aureus* were excluded from the new iteration because their previously estimated burden was minimal and estimates were available only for high-income countries.

### Countries and regions

National-level burden estimates were generated for all 194 WHO Member States, excluding overseas territories and disputed areas. For modelling and reporting purposes, Member States were clustered according to the six WHO regions, which were further stratified into seventeen subregions, based on the World Bank income classification (Low-income countries: ≤ $1,085;

Lower middle-income countries: $1,086 - $4,255; Upper middle-income countries: $4,256 - $13,205; High-income countries: $13,205+) (Table 1). We adopted the gross national income per capita calculated using the World Bank FY24 income classification, using 2022 data [16].

**Table 1.** Subregional clusters for WHO estimates of the global burden of foodborne diseases 2026 edition.

| WHO regions | Subregional clusters* | Countries |
| --- | --- | --- |
| African Region (AFR) | AB | Botswana, Equatorial Guinea, Gabon, Mauritius, Namibia, Seychelles, South Africa |
|  | C | Algeria, Angola, Benin, Cameroon, Cabo Verde, Comoros, Congo, Côte d'Ivoire, Eswatini, Ghana, Guinea, Kenya, Lesotho, Mauritania, Nigeria, São Tomé and Príncipe, Senegal, United Republic of Tanzania, Zambia, Zimbabwe |
|  | D | Burkina Faso, Burundi, Central African Republic, Chad, Democratic Republic of Congo, Eritrea, Ethiopia, Gambia, Guinea-Bissau, Liberia, Madagascar, Malawi, Mali, Mozambique, Niger, Rwanda, Sierra Leone, South Sudan, Togo, Uganda |
| Region of the Americas (AMR) | A | Antigua and Barbuda, Bahamas, Barbados, Canada, Chile, Guyana, Panama, Saint Kitts and Nevis, Trinidad and Tobago, United States of America, Uruguay |
|  | B | Argentina, Belize, Brazil, Colombia, Costa Rica, Cuba, Dominica, Dominican Republic, Ecuador, El Salvador, Grenada, Guatemala, Jamaica, Mexico, Paraguay, Peru, Saint Lucia, Saint Vincent and the Grenadines, Suriname |
|  | C | Bolivia (Plurinational State of), Haiti, Honduras, Nicaragua, Venezuela (Bolivarian Republic of) |
| South-East Asia Region (SEAR) | B | Indonesia, Maldives, Thailand |
|  | CD | Bangladesh, Bhutan, India, Myanmar, Nepal, Democratic People's Republic of Korea, Sri Lanka, Timor-Leste |
| European Region (EUR) | A | Andorra, Austria, Belgium, Croatia, Cyprus, Czechia, Denmark, Estonia, Finland, France, Germany, Greece, Hungary, Iceland, Ireland, Israel, Italy, Latvia, Lithuania, Luxembourg, Malta, Monaco, Netherlands (Kingdom of the), Norway, Poland, Portugal, Romania, San Marino, Slovakia, Slovenia, Spain, Sweden, Switzerland, United Kingdom of Great Britain and Northern Ireland |
|  | B | Albania, Armenia, Azerbaijan, Belarus, Bosnia and Herzegovina, Bulgaria, Georgia, Kazakhstan, Montenegro, North Macedonia, Republic of Moldova, Russian Federation, Serbia, Türkiye, Turkmenistan |
|  | C | Kyrgyzstan, Tajikistan, Ukraine, Uzbekistan |
| Eastern Mediterranean Region (EMR) | A | Bahrain, Kuwait, Oman, Qatar, Saudi Arabia, United Arab Emirates |
|  | BC | Djibouti, Egypt, Iran (Islamic Republic of), Iraq, Jordan, Lebanon, Libya, Morocco, Pakistan, Tunisia |
|  | D | Afghanistan, Somalia, Sudan, Syrian Arab Republic, Yemen |
| Western Pacific Region (WPR) | A | Australia, Brunei Darussalam, Cook Islands, Japan, Nauru, New Zealand, Niue, Singapore, Republic of Korea |
|  | B | China, Fiji, Malaysia, Marshall Islands, Palau, Tonga, Tuvalu |
|  | C | Cambodia, Kiribati, Lao People's Democratic Republic, Micronesia (Federated States of), Mongolia, Papua New Guinea, Philippines, Samoa, Solomon Islands, Vanuatu, Viet Nam |

\*WHO regions (as of September 2023) were subdivided into 17 subregions based on World Bank income classifications.

### Reference period

Estimates were generated for the period 2000–2021. The end year of the reference period was defined in function of WHO Global Health Estimates (GHE) and Institute for Health Metrics and Evaluation (IHME) Global Burden of Disease (GBD) estimates available at the time of modelling [17–18]. Data were collected and modelled from 1990 onwards, to allow for more robust trend estimates from 2000 onwards.

## Data collection

### Systematic literature reviews

Systematic reviews of key epidemiological input parameters, such as disease incidence, prevalence, and case fatality, played an essential role in the burden calculation activities. To ensure consistency, reviews conducted by WHO-commissioned experts were carried out following a standardized organization and documentation process. In total, nineteen review teams were commissioned to gather the required data.

To increase transparency and minimize bias, the commissioned teams were asked to develop a protocol in line with the PRISMA-P guidelines (http://www.prisma-statement.org/documents/PRISMA-P-checklist.pdf). Whenever possible, protocols were registered in Prospero for full transparency (https://www.crd.york.ac.uk/prospero/). Furthermore, to ensure coherence, each protocol followed a minimum set of requirements, including the study period, no language exclusion and inclusion of essential databases (**S4 Appendix**).

To support the imputation process, and to allow for appropriate propagation of uncertainty, a standardized data extraction template was provided (**S5 Appendix**). In addition, a data collection handbook was developed to support the commissioned teams (**S6 Appendix**). The extraction template contained a set of required variables for the different parameters, with optional fields for elements not relevant to all hazards (e.g., type of diagnostic test used). Throughout the review process, Sciensano and a member of the relevant TF were available for clarifications, and, when required, meetings with the reviewing team were organized. The team at Sciensano also reviewed the data extraction regularly to ensure correct understanding and consistency.

### Global burden of disease envelopes

For certain hazards, the burden estimates were directly adopted from existing global burden initiatives, or calculated as an attributable fraction of existing outcome-specific burden envelopes (see also <u>Burden calculation</u>). Whenever possible, we used WHO/GHE burden envelopes (by country, year, age, sex; together with uncertainty estimates). If these were not available, we used the IHME GBD envelopes. Although the GBD study uses modelling methods that differ from the present framework, these estimates provide a comprehensive and internally consistent source of epidemiological information for outcomes that would otherwise lack a suitable global envelope.

### Population data

Population and life expectancy data by country, year, age and sex were sourced from the United Nations World Population Prospects 2024 revision [19].

## Meta-regression

Literature searches typically do not provide epidemiological parameter estimates for every year-country combination. Therefore, extrapolation or imputation models are essential to construct a complete dataset across space and time. Furthermore, these models allow smoothing the available estimates, as they reduce variability due to sampling error across studies.

In the context of the WHO 2015 edition estimates, various approaches for imputing missing incidence data at the country level were tested and evaluated [20]. This process highlighted several pitfalls associated with using explanatory covariates, including risks of overfitting and the arbitrariness in covariate selection. To ensure parsimony and transparency, we adopted a hierarchical meta-regression model as the default method for imputing missing country-level data.

### Hierarchical meta-regression model

The full model estimates epidemiological parameters, such as incidence, prevalence or case fatality, across spatial and temporal dimensions while allowing for adjustments based on additional study-level covariates (**S7 Appendix**). The model is specified as follows:

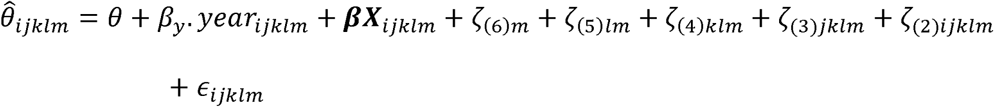

For data point *i*, study *j*, country *k*, subregion *l*, and region *m*, where:

- *θ̂*_*ijklm*_ is an estimate of the true effect size *θ*_ijklm_ for data point *i* nested in study *j*, nested in country *k*, nested in subregion *l*, nested in region *m*.
- *θ* is the global and universal intercept
- *β*_*y*_ is the regression coefficient for the global effect of year
- ***β*** is the regression coefficients vector for possible other study-level covariates ***X***
- _ζ(6)*m*_ is the random effect for region *m*
- _ζ(5)*lm*_ is the random effect for subregion *l*, nested in region m
- _ζ(4)*klm*_ is the random effect for country *k*, nested in subregion *l*, nested in region m
- _ζ(3)*jklm*_ is the random effect for study *j* nested in country *k*, nested in subregion l, nested in region *m*
- _ζ(2)*ijklm*_ is the random effect for data point i nested in study j, nested in country k, nested in subregion l, nested in region m
- *ε*_*ijklm*_ is the error term, pre-defined based on the study sample size.

The model was applied to log-transformed incidence, case fatality and mortality rates, and to logit-transformed prevalence ratios, population attributable fractions, and proportions.

The temporal trend, *β*_y_, was modelled as a global fixed effect, implying a shared time trend across countries and regions. Consequently, regional and country-level differences primarily arise from the hierarchical random effects structure and from differences in burden envelopes, rather than from region-specific temporal trends. This assumption was necessary due to data sparsity and lack of convergence in models allowing region-specific slopes.

Alternative temporal model specifications were explored, including random slope models allowing for region-specific time trends, as well as non-linear approaches. These models were compared with the linear time-trend specification using model fit statistics, convergence diagnostics, and the plausibility of the resulting estimates. However, they did not consistently improve model fit and were more prone to convergence issues, likely due to sparse and uneven temporal data coverage across hazards and regions. The linear global time trend was therefore retained as the most stable specification.

Depending on data availability, biological plausibility, and model convergence, alternative versions of the full model were applied across different hazards. For example, in some cases, only the regional effect was considered. In other cases, when most individual publications (studies) yielded only single data points, the random effect *<u>ζ</u>*_(2)ijklm_ for each data point was not considered. **S7 Appendix** provides an overview of the models used for the different meta-regressions.

### Bayesian implementation and convergence assessment

The models were implemented in a Bayesian framework to account for uncertainty arising from the estimation process. Specifically, we used the ‘brms’ package (version 2.22.0) in R (version 4.5.1), which interfaces with Stan [21]. We used flat priors for the regression coefficients, and weakly informative priors for the other coefficients — i.e., a Student-T(3) prior for the intercept, and half-Normal(0,1) priors for the standard deviations of the random effects. We fitted five chains, each consisting of 5000 iterations, of which the first 3000 were discarded as warmup. This resulted in 10,000 retained iterations for each quantity of interest. These parameters were treated as independent during the uncertainty propagation process.

Convergence was assessed by visually examining trace plots and ensuring Rhat statistics to be close to one. We also monitored divergent transitions and, when necessary, adjusted tree depth and delta values to improve sampling precision and model stability. To evaluate the robustness of the estimates to key modelling assumptions, sensitivity analyses were conducted by systematically varying modelling choices. These included alternative meta-regression specifications, such as models fitted on predefined subsets of the data and models incorporating additional covariates, to assess the influence of specific study characteristics. Additionally, alternative imputation hierarchies were explored by modifying the hierarchy used to inform imputation, alongside changes to the overall model specifications.Imputation process

After fitting the hierarchical meta-regression model to the available data, its posterior predictive distributions (i.e., the probability distributions derived from the fitted model parameters) were used to impute incidence values for countries with missing data. The imputation process followed a hierarchical structure:

- If data were available for other countries in the same subregion, the subregion-year-specific estimate was used.
- If the subregion lacked data but countries in the broader region had data, the region-year-specific estimate was used.
- If no data were available for the region, the global-year-specific estimate was applied.

For some hazards, specific countries were not included in the imputation model. In these cases, incidence, mortality, and burden were assigned a value of zero. For *Clonorchis sinensis*, *Opisthorchis viverrini* and *Opisthorchis felineus*, *Paragonimus*, intestinal flukes, *Fasciola* and *Fasciolopsis*, *Ascaris*, *Trichinella* and *Taenia solium*, this reflected the absence, or negligible occurrence, of locally acquired human cases attributable to foodborne transmission in those countries. For cholera, countries with no reported cases since 2015 were assumed to have no incidence. For EAEC and EPEC, data availability was deemed insufficient to produce reliable burden estimates. **S8 Appendix** provides the list of so-called “burden-free” countries for each hazard.

After imputation, and depending on the obtained estimates, the data were critically assessed to identify and explain any unexpected results. This involved checking whether the imputed estimates were consistent with the observed data, investigating studies that might have overly influenced the results, and consulting with TF members, and, where necessary, additional external experts, to evaluate estimate reliability. In cases where discrepancies were found, further evaluation was conducted to determine whether they stemmed from data limitations, model assumptions, or underlying epidemiological factors. Data adjustments (adding/removing data points, adjusting sample sizes), model changes (adding/removing covariates) and methodology adjustments for the imputation estimates were conducted and documented in collaboration with experts from each TF to achieve reliable estimates.

## Burden calculation

### Disease models

In the hazard and incidence-based approach, DALYs are obtained by adding up the YLD and YLL contributions of the different health states causally associated with the concerned hazard. These health states can be acute or chronic stages of the disease, can include specific sequelae, and death, as the ultimate sequela. A disease model, or outcome tree, aims to visualize these different health states, whereas a *computational* disease model maps the data and calculation processes leading to the estimated cases in each of the health states (Figure 2).

**Figure 2.**
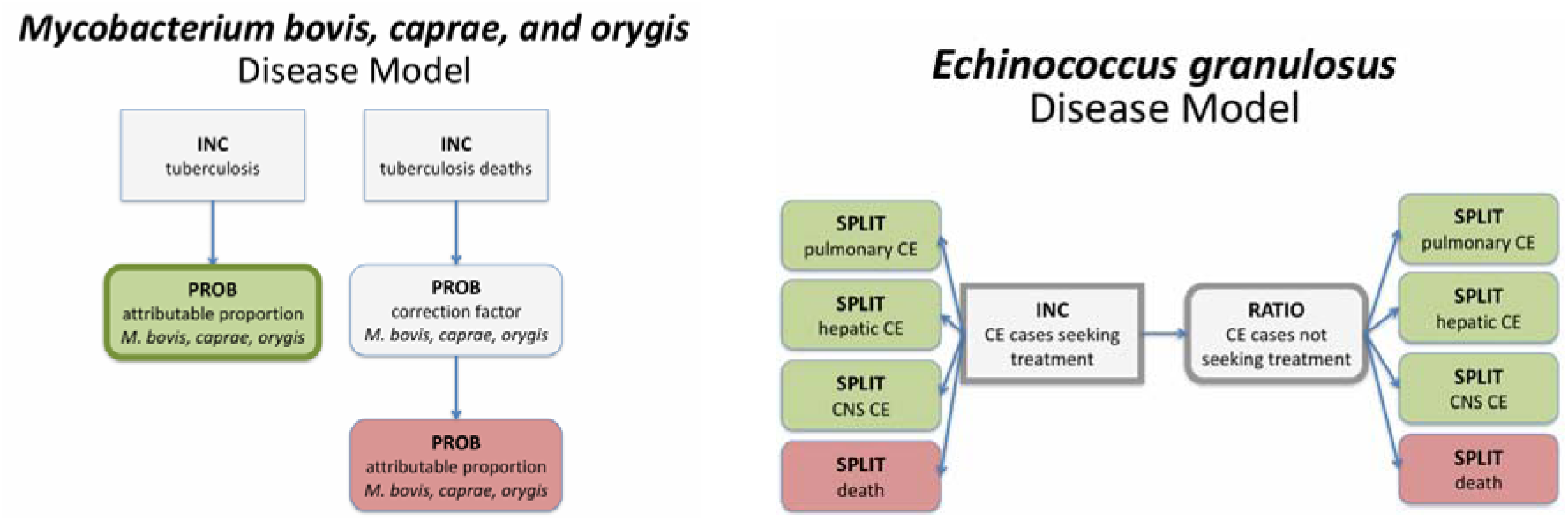
Examples of computational disease models. In addition to visualizing the included health states, they also visualize the flow of the calculations. Grey nodes do not contribute DALYs, green nodes contribute YLDs, red nodes contribute YLLs; nodes with a thick outline contribute incident cases. INC = incidence; PROB = probability; SPLIT = mutually exclusive fractions; CE = cystic echinococcosis; CNS = central nervous system.

A first key step towards a foodborne disease burden estimate is, therefore, the construction of a disease model. This model consists of four elements – i.e., the nature of the health states, the calculation process to obtain health state incidence, the duration, and the disability weight of each health state. **S9 Appendix** provides an overview of the disease models applied per hazard.

### Hazard-attributable health state incidence

In general, three main approaches can be distinguished for calculating hazard-attributable health state incidence:

- **Direct estimation approach**: the health state incidence is directly available; e.g., laboratory-based surveillance systems provide estimates of the incidence of *Salmonella*-attributable diarrhoea; other examples include a direct evaluation of peanut allergy incidence, or a direct evaluation of trichinellosis incidence.
- **Attribution approach (a.k.a, syndrome or population data scaled-down)**: the incidence of the symptom or syndrome is multiplied with the attributable proportion for a given hazard; e.g., the overall diarrhoea incidence may be multiplied with the *Salmonella* etiological fraction (proportion of cases attributable to *Salmonella*); other examples include the attribution of a liver cancer envelope to aflatoxin based on Population Attributable Fractions (PAF), or the attribution of an epilepsy incidence envelope to neurocysticercosis based on categorical attribution.
- **Transition approach**: the incidence of infection with, or exposure to, the hazard is multiplied with the probability of developing a given symptom; e.g., the incidence of *Salmonella* infections may be multiplied with the probability of developing diarrhoea given *Salmonella* infection; other examples include the evaluation of congenital toxoplasmosis symptoms as a fraction of all seroconversions, or the risk assessment approach to estimate infertility incidence from dioxin exposure.

These estimates aim to reflect the true, population-level incidence. Whenever needed, incidence estimates were therefore corrected for underestimation, as an additional component in the computational disease model.

### DALY methods

#### Standard expected years of life lost

The remaining life expectancy table (standard loss function) to calculate years of life lost by age, was adopted from the WHO/GHE 2021 revision [17]. The WHO/GHE utilizes the frontier national life expectancy derived from the lowest projected age-specific mortality rates for the year 2050 by the World Population Prospects 2024. The highest projected life expectancies for the year 2050 are projected to be achieved with a life expectancy at birth of 92 years.

#### Disability weights

We adopted the disability weights from Salomon et al. (2015), [22], as used in the most recent iterations of the Global Burden of Disease study. When the relevant health state was not available in the WHO/GHE list, proxy health states were selected or alternative sources consulted such as the European Disability Weights study [23]. These sources are compatible with the WHO/GHE list and include additional health states for infectious diseases.

#### Duration of non-fatal health states

The duration until remission or death is required for each of the non-fatal health states. These durations can differ by age, sex, region, depending on clinical evidence and available data. The disease durations from the previous editions were generally retained [4], unless new evidence was identified that supported an update. For lifelong sequelae, the national age and sex-specific life expectancy was used as duration, as this reflects the expected time an individual lives with the condition.

#### Social weighting

As for the previous iteration, and in line with WHO/GHE [17], no age weighting or time discounting were applied.

#### Comorbidity adjustments

As for the 2015 edition, comorbidity adjustments were not performed in a systematic way, mainly because only a subset of causes of ill health was considered, in contrast to the Global Burden of Disease Study. However, specific considerations were made for invasive non-typhoidal salmonellosis and Human Immunodeficiency Virus (HIV), where the burden of joint infections was attributed to HIV.

### Probabilistic burden assessment

The DALY calculations were implemented in a probabilistic framework, using 1000 Monte Carlo simulations (or 500 for the hazards where IHME/GBD envelopes were used). The analyses were implemented in R, with the code of the computational framework made available as an open access R package ‘dalymod’, distributed via GitHub (https://github.com/brechtdv/dalymod/).

The calculation process yielded estimates of incidence, mortality, YLD, YLL, and DALY, by hazard, health state, age, sex, country, and year. Estimates are produced as absolute numbers, rates per 100,000 population, and rates per case. Probability distributions were summarized by their mean and 95% uncertainty interval. Regional, global and hazard totals were obtained by aggregating country-level estimates within each Monte Carlo iteration. This approach preserved the full uncertainty structure during aggregation, rather than summing only point estimates.

## Source attribution

The main aim was to quantify the disease burden resulting from foodborne exposure to potentially foodborne hazards. However, many of the hazards considered are not transmitted solely by food, but have several potential transmission pathways (e.g., contact with animals, human-to-human, water, soil, or other). For these hazards, it was necessary to attribute a proportion of their overall burden to foodborne exposure.

Some hazards were considered 100% foodborne, i.e., *Taenia solium*, *Clonorchis sinensis*, minute intestinal flukes, *Opisthorchis* spp., *Paragonimus* spp., *Listeria monocytogenes*, *Clostridium botulinum*, *Trichinella,* Cassava cyanide, and peanut allergen. In addition, FERG estimated the foodborne disease burden for five chemical hazards (aflatoxin B1, inorganic arsenic, cadmium, dioxins, and methylmercury) for which attribution is embedded in the disease burden estimation, which relies on exposure assessment through food sources within each population.

For the remaining hazards, estimates were retrieved from a structured expert judgement study using Cooke’s Classical Model commissioned to a team of specialists to estimate the proportion of human disease burden attributable to different transmission pathways and to specific foods within the foodborne pathway, thereby generating hazard-specific attribution estimates for each subregion. Experts’ individual assessments were obtained for a given hazard/subregion via elicitation interviews and mathematically aggregated using a validated framework. When expert data were not sufficient for a given subregion, regional, economical or global proxies were considered; for more details, see [13]. Resulting aggregated distribution functions were obtained for each hazard/subregion [24]. Consequently, this study yielded probabilistic estimates of the proportion of incident cases that are foodborne, expressed as empirical cumulative distribution functions from which random samples could be drawn. Foodborne cases, deaths, YLDs, YLLs and DALYs were then obtained by multiplying the vectors of random values for these parameters with a vector of random values for the foodborne proportion.

## Governance and transparency

Throughout the process, different steps were built in to ensure internal consistency and quality of the estimates. Systematic reviews were conducted according to common standards, and regular follow-up was foreseen to ensure completeness and correctness of data extraction. Meta-regression and burden estimates were critically reviewed by the hazard group-specific TFs, leading to further refinements in data management and modelling strategies. In addition, a whole of FERG review ensured global plausibility of the estimates. Finally, the WHO country consultation process provided an opportunity for country-specific feedback on the estimates, including the provision of novel data.

Transparency of the process was guaranteed by adhering to the standards of reproducible research, including the development of open source and open access R packages for the computational framework (dalymod) and the study-specific settings and input parameters (FERG2), as well as the dissemination of outputs as reproducible R Markdown files via GitHub.

The final estimates are available through the WHO Global Health Observatory (https://www.who.int/data/gho/data/themes/topics/foodborne-diseases-estimates) and an accompanying dashboard designed for detailed exploration and comparison (https://www.who.int/teams/nutrition-and-food-safety/monitoring-nutritional-status-and-food-safety-and-events/foodborne-disease-estimates/2nd-edition-(2026)). Scientific publications will present the methods, results, and limitations of the individual hazard estimates.

## Discussion

This manuscript describes the methodology adopted for generating the WHO 2026 edition estimates of the global, regional and national burden of foodborne disease. Compared to the WHO 2015 edition estimates, this framework provides a series of significant methodological and practical improvements. First, the scope was expanded from 31 to 42 hazards. Second, estimates were generated annually for the period 2000–2021, rather than for a single reference year. Third, the framework produced country-level estimates for all WHO Member States, in addition to presenting only regional and global estimates. Fourth, the use of a harmonized hierarchical modelling approach improved internal consistency across hazards and countries. Finally, the updated framework strengthened uncertainty propagation, reproducibility, and transparency through Bayesian modelling, Monte Carlo simulation, and open source computational workflows.

The WHO 2026 edition estimates of the global burden of foodborne diseases provide a complementary set of estimates to the GBD study, which is the biggest global health indicator development effort to date [25]. The WHO 2026 edition estimates include a series of hazards that are not (explicitly) included in the GBD estimates, such as *Toxoplasma gondii* and arsenic. Furthermore, the consistent use of a hazard- and incidence-based approach also distinguishes the WHO estimates from the GBD study. This distinction is particularly important for diarrhoeal hazards such as *Campylobacter* and *Salmonella*, which the GBD study considers only as causes of diarrhoeal disease, thereby excluding the burden associated with sequelae such as Guillain–Barré syndrome. Finally, GBD does not attribute disease burden specifically to foodborne transmission. These factors, together with broader differences in modelling approaches, imply that the estimates are not directly comparable.

Even though presenting a significant advance, the WHO 2026 edition modelling framework is subject to a series of limitations. The updated framework represents a complex undertaking involving multiple hazards, heterogeneous data sources, and an extensive international collaboration network. This complexity introduced a need for hazard-specific and often ad hoc approaches to modelling, extrapolation, and imputation, making it difficult to maintain a fully standardised analytical framework across hazards. These challenges were particularly evident for parasites, which often display highly focal transmission patterns within and between countries. In addition, varying proportions of symptomatic and asymptomatic infections affect detectability in surveillance systems and contribute to under-ascertainment. For chemical hazards, burden estimates relied more heavily on (comparative) risk assessment approaches, which are inherently more uncertain. Addressing these challenges required advanced statistical methods, robust computational infrastructure, transparent documentation and version control, and continued engagement with data providers and national stakeholders.

Despite substantial improvements in data availability compared with the previous edition, important data gaps remained (**S8 Appendix**), and available epidemiological data were highly heterogeneous with respect to study design, diagnostic methods, case definitions, and geographical coverage. Although standardized systematic review protocols were implemented to enhance consistency, residual bias due to underreporting, misclassification, and publication bias could not be excluded. Prevailing data gaps resulted in a strong reliance on imputation, in particular in low- and middle-income regions, where the burden is expected to be highest. Data limitations furthermore led to the impossibility of generating more specific time trends (e.g., showing different temporal evolutions by country), or more detailed age and sex distributions. The latter also precluded the calculation of age-standardised rates. Finally, data scarcity and heterogeneity also resulted in wide uncertainty intervals in the final DALY estimates for several hazards. Future studies based on nationally representative data would help narrow these intervals and improve the precision of global burden estimates for these pathogens.

The framework also does not claim to be exhaustive, and could in the future be expanded to include additional (emerging) foodborne hazards, such as pesticides, *Angiostrongylus* spp., or Hepatitis E Virus. The current framework does not explicitly account for major global drivers such as the COVID-19 pandemic, antimicrobial resistance, or climate change. These factors may influence foodborne disease transmission dynamics, healthcare access, and outcomes.

Despite the acknowledged limitations, the WHO 2026 edition modelling framework underpins the estimation of the global burden of foodborne diseases by enabling more detailed and up-to-date estimates through a rigorous and robust approach. The revised framework also provides a basis for routine monitoring of foodborne disease burden, allowing for the assessment of progress towards global food safety targets and other objectives. Additionally, the identified data gaps have the potential to motivate national authorities to enhance data collection systems through national surveillance systems or targeted studies, while the inherent limitations of a global modelling effort may stimulate interest in conducting national-level burden studies on foodborne diseases.

## Conclusion

This manuscript describes the updated computational framework adopted by WHO for estimating the 2026 edition of the global burden of foodborne diseases. Despite ongoing data limitations and modelling constraints, the WHO 2026 edition estimates represent the most comprehensive, transparent, and reproducible assessment to date. The results are expected to strengthen global and national food safety monitoring, identify critical data gaps, and guide future investments in surveillance and hazard-specific studies.

## Declarations

### Ethics approval and consent to participate

Not applicable.

## Consent for publication

Not applicable.

### Availability of data and materials

The code of the computational framework is available as an open access R package ‘dalymod’, available via https://github.com/brechtdv/dalymod. The study-specific settings and input parameters are available as an open access R package ‘FERG2’, available via https://github.com/brechtdv/FERG2. The model outputs are available as reproducible R Markdown files via https://github.com/fbdburden.

## Author contributions

Conceptualization, Methodology: All

Data curation, Formal analysis: BD, CDB, LV, CF, KF Funding acquisition: EB, YM, RK

Project administration, Supervision: BD, YM, MK Writing – Original Draft Preparation: BD, CDB, LV Writing – Review & Editing: All *Acknowledgments*

Not applicable.

## Supporting Information

**S1 Appendix. STROBOD checklist.**

**S2 Appendix. GATHER checklist.**

**S3 Appendix. Hazard prioritization criteria.**

**S4 Appendix. Concept note for the WHO call for expressions of interests on conducting systematic reviews and other studies for estimating the burden of foodborne diseases.**

**S5 Appendix. Data extraction template.**

**S6 Appendix. Data collection handbook.**

**S7 Appendix. Burden approaches and models fitted for each input parameter.**

**S8 Appendix. Summary of data points, studies and blocked countries by country and hazard.**

**S9 Appendix. Disease models.**

