## Supplementary material for "Computational framework for the World Health Organization estimates of the global, regional and national burden of foodborne diseases 2026 edition": S1 Appendix

| Item number | Domains and description of the recommended items | Reported on page number |
|---|---|---|
| **TITLE** | | |
| 1 | Identify the study as a burden of disease assessment by including keywords (e.g., Years of Life Lost, Years Lost due to Disability, Disability-Adjusted Life Years, burden of disease etc), and describe the study setting. | 1 |
| **ABSTRACT** | | |
| 2 | Provide a summary of objectives, study setting, methods (including data sources and key methodological design choices used), results (including point estimates and, if applicable, uncertainty intervals), and conclusions. | 2-3 |
| **INTRODUCTION** | | |
| 3 | Present background information to the study, its study aim(s), and its relevance for health policy or practice. | 4-5 |
| **METHODS** | | |
| **Study setting** | | |
| 4 | Report for which cause(s) the burden was calculated. Provide a case definition, e.g., in terms of an internationally recognized classification system such as the International Classification of Diseases and Related Health Problems 10th Revision. | 6-7 |
| 5 | Report the reference population and any stratification of the reference population for the burden of disease assessment, i.e., the population for which the burden was calculated. This may include the geographical location (e.g., country or province/state), and whether the general population or a specific subset of the population (e.g., females, adolescents aged 10-19 years, etc) was considered. | 7-9 |
| 6 | Report the reference time period (e.g., year(s), month(s)) of the study. This refers to the time period to which the burden of disease estimates refer. | 9 |
| **Epidemiological and demographic input data** | | |
| 7 | Report the sources, values, ranges, and, if used, probability distributions for all epidemiological input parameters. Report reasons or sources for distributions used to represent uncertainty where appropriate. Providing a (supplementary) table to show all epidemiological input parameters and respective sources and assumptions is strongly recommended. | 20 |
| 8 | Describe all possible data manipulations, such as bias corrections, data integration steps, or methods to ensure internal consistency of the data inputs. | 11-15 |

| Item number | Domains and description of the recommended items | Reported on page number |
|---|---|---|
| 9 | Report the sources and values of any population data used. If applicable, report the standard population used to calculate age-standardized rates. | 11 |
| **DALY methods** | | |
| 10 | Report the age-conditional life expectancy used for calculating Years of Life Lost (i.e., national, regional, or aspirational life tables) or other methods (e.g., potential years of life lost, proportion of premature deaths under a selected age threshold etc). | 17 |
| 11 | Report the perspective taken for calculating Years Lost due to Disability, i.e., incidence or prevalence perspective. | 5 |
| **Disease model** | | |
| 12 | Describe the disease model. Present and justify the included health outcomes and health states. Providing a (supplementary) figure visualizing the disease model is strongly recommended. | 15-17 |
| 13 | Report the source(s) and values of the used disability weights. Providing a (supplementary) table depicting the health states, brief lay descriptions, and the numerical values followed by its uncertainty intervals is strongly recommended. | 17 |
| 14 | If new disability weights were elicited, provide information on how the health states were described and the elicitation procedures. As a minimum to the latter, describe which valuation technique was used and which reference group and size of the group (also known as panel of judges) evaluated the health states. Providing a supplementary table with a description of the valuation technique and brief lay descriptions used is strongly recommended. | N/A |
| 15 | Report the source(s) and values of the used durations (if applicable). Providing a (supplementary) table depicting the health states and the numerical values followed by its uncertainty intervals is strongly recommended. | 17-18 |
| 16 | Report the source(s) and values of the used conditional probabilities, severity distribution, and/or transition rates. Providing a (supplementary) table depicting the parent/child health outcomes and health states and the numerical values followed by its uncertainty intervals is strongly recommended. | Ref 8-12 |
| **Multimorbidity adjustments** | | |
| 17 | Report whether or not multimorbidity adjustments were applied to any of the input variables in the estimation of Years Lost due to Disability. If applied, describe which multimorbidity adjustment method was used. | 18 |
| **Social weighting factors** | | |
| 18 | Report whether or not age weighting was applied. If applied, describe which parameters were used. | 18 |

| Item number | Domains and description of the recommended items | Reported on page number |
|---|---|---|
| 19 | Report whether or not time discounting was applied. If applied, describe which discount rate was used. | 18 |
| **Uncertainty and scenario analysis** | | |
| 20 | Describe any methods used to perform uncertainty and variable importance (sensitivity) analyses. If, for example, Monte Carlo simulations were used, report the number of iterations. | 18 |
| 21 | Describe any scenario analyses that were performed. Present the rationale and the alternative data inputs defining the alternative scenarios. | N/A |
| **RESULTS** | | |
| 22 | Report the point estimates and, if applicable, the uncertainty interval of the burden of disease estimates. Provide both absolute values, crude rates (optional), and age-standardized rates per 100,000 in a table or figure. | Ref 8-14 |
| 23 | If applicable, report the results of the scenario analyses. Tables and/or figures illustrating findings on the scenario analyses are strongly recommended. | N/A |
| **DISCUSSION** | | |
| 24 | Summarise the key study findings and describe how they support the conclusions reached. | Ref 8-14 |
| 25 | Discuss how the findings fit within current knowledge. Discuss potential implications for public health practice. Compare the results with those of other studies, and discuss methodological design differences, if relevant. | Ref 8-14 |
| 26 | Discuss strengths and limitations, and the generalisability of the study findings. If applicable, discuss the results of the uncertainty and scenario analyses. | Ref 8-14 |
| **OPEN SCIENCE** | | |
| 27 | Make the source code or computational model(s) available as supporting information or via a dedicated open access repository (e.g., GitHub). | 24 |
| 28 | Describe how the study was funded and the role of the funder in the identification, design, conduct, and reporting of the analysis. Describe other non-monetary sources of support or any potential conflict(s) of interest of the study contributor(s) in accordance with the journal policy. | 24 |
