## Supplementary material for "Computational framework for the World Health Organization estimates of the global, regional and national burden of foodborne diseases 2026 edition": S3 Appendix

### S1 Appendix. Hazard prioritization criteria.

1. Estimated frequency of occurrence
    a. High: the hazard occurs frequently and has a (near) worldwide distribution.
    b. Medium: the hazard occurs frequently but has a focalized distribution.
    c. Low: the hazard occurs infrequently, or has a limited geographical distribution.
2. Estimated severity of cases
    a. High: the hazard is associated with severe outcomes and/or death, which occur in a significant number of cases.
    b. Medium: the hazard is associated with severe outcomes and/or death, but these occur in a minority of cases.
    c. Low: the hazard generally leads to mild and asymptomatic cases.
3. Estimated economic impact of hazard (e.g. due to livestock productivity losses or trade impacts)
    a. High: the hazard is associated with major economic impact in a majority of countries.
    b. Medium: the hazard is associated with significant economic impact in at least a considerable number of countries.
    c. Low: the hazard is associated with little to no economic impact.
4. Estimated data availability
    a. High: data are readily available for a sufficiently large number of countries, spanning the different WHO regions. Imputation will only have a moderate effect on the final estimates.
    b. Medium: data are readily available, but without complete global coverage. Imputation will have a major effect on the final estimates.
    c. Low: data are not readily available, or only for a limited part of the world. Imputation is not deemed feasible.
5. Estimated proportion foodborne
    a. High: the hazard is only transmitted via food
    b. Medium: the hazard is transmitted via food in a majority of cases
    c. Low: the hazard is transmitted via food in a minority of cases
6. Inclusion in FERG1
    a. Yes
    b. No
