## Supplementary material for "Computational framework for the World Health Organization estimates of the global, regional and national burden of foodborne diseases 2026 edition": S4 Appendix

### WHO call for expressions of interests – conducting systematic reviews and other studies for estimating the burden of foodborne diseases

Concept Note

**Version 4 October 2022**

#### Introduction

The WHO Foodborne Disease Burden Epidemiology Reference Group (FERG)[1] has been reconvened in May 2021 to update the global burden of foodborne disease estimates, which were previously published in 2015.[2] The updated WHO global report is requested by Member States by 2025.

In order to prepare the updated estimates, WHO wishes to commission a number of pieces of work, principally systematic reviews of scientific and other literatures. This Concept Note describes the overall work programme, and methodology. Details for specific work items will be described in separately published Terms of Reference.

#### Background

Foodborne diseases are a major cause of human morbidity and mortality. According to the 2015 WHO estimates calculated with the FERG from 2007 to 2015, foodborne diseases caused 600 million illnesses, 420 000 deaths, and 33 million disability-adjusted life years (DALYs) in 2010. Foodborne diseases disproportionately and adversely impact children. According to these estimates, although children <5 years of age represent only 9% of the global population, 40% of the foodborne disease burden is borne by children in this age group. There are also considerable differences in the burden of foodborne diseases among sub-regions with the highest burden per population observed in Africa.

In 2020, the Seventy-third World Health Assembly adopted the resolution, *"Strengthening efforts on food safety"* (WHA73.5[3]), requesting WHO to monitor regularly, and to report to Member States on, the global burden of foodborne and zoonotic diseases at national, regional and international levels, and in particular to prepare, by 2025, an updated report on the global burden of foodborne diseases with up-to-date estimates of global foodborne disease incidence, mortality and disease burden in terms of DALYs. Following the re-establishment of the FERG in 2021, WHO has conducted three expert meetings to date in July 2021, October

[1] https://www.who.int/groups/foodborne-disease-burden-epidemiology-reference-group-(ferg)
[2] https://apps.who.int/iris/handle/10665/199350
[3] https://apps.who.int/gb/ebwha/pdf_files/WHA73/A73_R5-en.pdf

2021, and April 2022, respectively. Meeting details have been published outlining the proceedings and outcomes.[4] [5] [6] The fourth expert meeting is planned in November 2022 in Geneva, Switzerland.

#### Objectives and of the concept note

This Concept Note covers the work to be commissioned for three objectives:

1) To provide data and information to support the estimation of the burden of foodborne diseases from specified hazards
2) To provide data and information to contribute with the monitoring activities on the implementation and impact of the Global Food Safety Strategy[7]
3) To provide data and information to enable FERG to support foodborne disease studies in individual countries

#### Methodology

Systematic reviews play an essential role in the FERG activities, and much of the commissioned work will be of this type. As the systematic reviews will be conducted by a variety of (commissioned) experts, it is important to standardize the organization and documentation of the systematic reviews. This Concept Note provides an overview of the protocol for systematic reviews. Exact details on each review are published in respective Terms of Reference.

##### Protocol for all systematic reviews

The protocol for the systematic reviews must be developed in line with the PRISMA-P guidelines (http://www.prisma-statement.org/documents/PRISMA-P-checklist.pdf). For full transparency, protocols must be registered in Prospero (https://www.crd.york.ac.uk/prospero/).

For consistency, the protocol should follow a minimum set of requirements:

1) Databases
    i. Systematic searches are required to consider both peer-reviewed and grey literature
    ii. Peer-reviewed literature needs to be searched for in at least the following repositories: PubMed, Web of Science, Embase, Scopus, and INASP Journals Online project.
    iii. Grey literature needs to be searched for in at least the following repositories: IRIS (WHO), Oaister, Google Scholar
    iv. Where relevant, national surveillance data can be collected
2) Language requirements
    i. Systematic searches are required to include both English and non-English papers
3) Data period
    i. Data need to be collected from studies published between 1990 and the most recent time period

[4] First meeting of the Foodborne Disease Burden Epidemiology Reference Group (FERG) 2021-2024 (who.int)
[5] Second meeting of the Foodborne Burden Disease Epidemiology Reference Group (FERG2) 2021-2024 (who.int)
[6] Third meeting of the Foodborne Burden Disease Epidemiology Reference Group (FERG2) 2021-2024 (who.int)
[7] https://apps.who.int/gb/ebwha/pdf_files/EB150/B150_25-en.pdf

#### Protocol for data extraction for systematic reviews of epidemiological parameters

Data we are collecting can roughly be classified into four types (refer to generic outcome trees):

i. Incidence rates
ii. Probabilities or proportions
iii. Age and sex distribution of cases and deaths (can be region-specific)
iv. Duration of health states (can be region-specific)

A data extraction table is required for each epidemiological parameter, e.g., incidence, prevalence, proportion, duration. To support and standardize the imputation process, and to allow for appropriate propagation of uncertainty, a standardized data extraction template is provided. This comprises the following variables:

| | |
|---|---|
| **SOURCE_ID** | Identification number of the input source |
| **SOURCE_AUTHOR** | First author of input source |
| **SOURCE_YEAR** | Year of publication |
| **SOURCE_TITLE** | Title of source |
| **SOURCE_DOI** | DOI of source (if available) |
| **SOURCE_URL** | URL of source, other than DOI (if available) |
| **REF_YEAR_START** | Starting year of the data derived from the input source |
| **REF_YEAR_END** | End year of the data derived from the input source |
| **REF_LOCATION** | Geographic location for which the input source was used |
| **REF_AGE_START** | Numerical value of starting age of the population of the data derived from the input source |
| **REF_AGE_END** | Numerical value of ending age of the population of the data derived from the input source |
| **REF_SEX** | Sex of the population of the data derived from the input source |
| **REF_SAMPLE_SIZE** | Sample size of data derived from input source |
| **VALUE_MEAN** | Mean estimate of the data derived from the input source |
| **VALUE_MEDIAN** | Median estimate of the data derived from the input source |
| **VALUE_SE** | Standard error of the data derived from the input source |
| **VALUE_P0** | 0th percentile (minimum) of the data derived from the input source |
| **VALUE_P2_5** | 2.5th percentile of the data derived from the input source |
| **VALUE_P5** | 5th percentile of the data derived from the input source |
| **VALUE_P10** | 10th percentile of the data derived from the input source |
| **VALUE_P25** | 25th percentile of the data derived from the input source |
| **VALUE_P75** | 75th percentile of the data derived from the input source |
| **VALUE_P90** | 90th percentile of the data derived from the input source |
| **VALUE_P95** | 95th percentile of the data derived from the input source |
| **VALUE_P97_5** | 97.5th percentile of the data derived from the input source |
| **VALUE_P100** | 100th percentile (maximum) of the data derived from the input source |
| **VALUE_X** | Number of events in data derived from input source |
| **VALUE_N** | Sample size of data derived from input source |

**Instructions:**

1) A data extraction table needs to be made for each epidemiological parameter, such as incidence, prevalence, proportion, duration. Please copy data sheets as necessary.
2) The data extraction table needs to be filled in a consistent way, using coherent names and spelling for the different entries. Numerical fields should only include numerical values, no text. Missing values should be kept as blank cells.
3) If a single paper provides multiple breakdowns, the most detailed breakdowns need to be provided as separate rows in the table (thus duplicating the source identifier variables).
4) Additional variables (columns), such as the diagnostic method, can be added if deemed relevant.
5) Not all "VALUE" variables need to be filled in, only those that are directly obtained from the source.

#### Overall timeline (indicative)

**Start date:** 30 September 2022 **End date:** December 2023

#### Deliverables and timeline for delivery

The contractors will deliver a final dataset and report documenting results, to be further analyzed and interpreted by WHO and the FERG, especially the respective taskforces and the computational taskforce. It is expected that this review will result in a manuscript for publication in at least one peer-review journal, that must adhere to the WHO policy on Open Access[8]. Contractors are to lead the writing process, in close coordination with the relevant taskforces, respectively. The publication process will be governed by the existing publication policy, and authorship is subject to the recommendations for defining the role of authors and contributors published by the International Committee of Medical Journal Editors (ICMJE).

#### Other requirements

The study team or individual will be selected from the submitted expressions of interest and based on the qualifications and skills (see specifications in relevant section). Geographical and gender diversity is encouraged for applications from teams. Scientists will participate in their individual capacity rather than as a representative of their employer. Once shortlisted, each individual or team member will also need to complete the standard WHO Declaration of Interest form, which will be assessed for conflict of interests. The individual or team leader may be asked to further elaborate the expression of interest in a virtual video meeting with the WHO Secretariat. The final candidates will be selected through a competitive process in accordance with WHO's policies and procedures.

[8] https://www.who.int/about/policies/publishing/open-access

### HOW TO APPLY FOR A CALL FOR EXPRESSIONS OF INTEREST

#### Please note the following requirements before proceeding with your application.

To complete an application, an applicant (the team leader if it is more than a person) must provide responses to questions explicitly detailed in the application portal linked below. It is important to have all information prepared prior to applying online, as it is not possible to return to the portal to modify your submission:

1) Reference number (found in the Terms of Reference)
2) Contact information from the main focal point only (such as the Lead Investigator)
3) Cover letter/statement of motivation, including a maximum of 600 words detailing why your team are submitting this Expression of Interest, and why you believe your team is the most suitable to undertake this work. **It is recommended to prepare this in a separate Word document so you can copy and paste text into the application.**
4) Proposed fee for undertaking the work (in USD)
5) Ideal start date and completion dates to undertake the work
6) ONE document (ideally in PDF format) that includes every Curriculum Vitae (CV) of the proposed research team or an individual applicant.
7) ONE document (ideally in PDF format) that includes a brief biography of an applicant or each research team member (max 150 words per person).

#### Apply here: **https://extranet.who.int/dataformv3/index.php/998268?lang=en**

Contact: WHO secretariat
