## Supplementary material for "Computational framework for the World Health Organization estimates of the global, regional and national burden of foodborne diseases 2026 edition": S6 Appendix

### HANDBOOK DATA EXTRACTION FERG

*Last updated: 30.08.2023*

This handbook describes the different elements of the Excel template for data extraction provided by the Sciensano team. If you have any questions about the template or handbook, please feel free to contact us at.

The excel template contains the following elements:

- Data extraction templates for different epidemiological parameters
- A codebook describing the variables to be extracted

#### NAMING THE DATA EXTRACTION WORKBOOK

For consistency, please use the following format for naming your data extraction workbook:

"**ferg2-*Task force-Hazard-YYYYMMDD***"

… *Task force*: edtf, pdtf, cttf or satf.
… *Hazard*: pathogen, toxin or other hazard under investigation.
… *YYYYMMDD*: refers to the date on which you send the data extraction table to the Sciensano team so we can keep track of different versions.

Please only use lower case letters and hyphens ("-").

Example: ferg2-pdtf-trichinella-20230301

#### DATA EXTRACTION SHEETS

A separate data extraction table (i.e. excel sheet) needs to be used **for each epidemiological parameter.**

Sheets for the following parameters are predefined in the template:

- Incidence
- Prevalence
- Mortality
- Duration
- Sequelae

Depending on the deliverables agreed upon between your team and WHO, either all of them or only a subset needs to be used. Please contact the Sciensano team if you need another template for a different type of epidemiological parameter.

#### VARIABLES

##### VARIABLE TYPES

Data is collected on four different types of variables:

| ***Variable Type*** | ***Name*** | ***Description*** |
|---|---|---|
| *Source identifier* | SOURCE_ | General information about the input source |

| | | |
|---|---|---|
| *Reference* | REF_ | Specifics about the study location, population & time |
| *Value* | VALUE_ | Variables concerning the output estimates. Not all need to be filled in, only those that are directly obtained from the source. |
| *Optional* | OPT_ | Optional variables that may be used as the reviewers deems fit. We provided some examples in the templates but more can be added if needed. |

The variable names for the source, reference and value variables **must not** be changed.

If a single input source provides multiple breakdowns, e.g. for different years or by sex, the most detailed breakdowns need to be extracted **as separate rows** in the table as well as the general one. The source identifier variables are duplicated across these rows. The breakdown level is specified in the reference variables and the corresponding results are entered in the value variables.

#### DATA TYPES

Each variable has one of the following data types:

| ***Data type*** | ***Description*** |
|---|---|
| *String* | Free text; please use consistent spelling across rows wherever possible. |
| *Integer* | Only enter whole number values; no text. |
| *Numerical* | Only enter numerical values, including decimal values; no text. |
| *Factor* | Chose from predefined drop-down options.<br>*Attention: The source values for the drop-down options are specified in a hidden sheet and cannot be modified. If an option is missing, you may either select "Other" and specify in the REF_NOTES column, or – especially if the option is needed more often – you can contact us to include it in the template.* |

#### DELETING AND ADDING VARIABLES

Deleting variables:

If any of the variables starting with **"OPT_"** are not relevant to your review, **you may delete them** from the templates. Please do not delete any of the other variables.

*Special case: for commissioned work other than systematic reviews, also other types of variable may be omitted, depending on the purpose of the data extraction. If this applies to your work,* ***please send your adjusted template to the Sciensano team before starting with the data extraction*** *to avoid problems with missing information later on.*

Adding variables:

If you want to extract information on variables not predefined in the templates, **you can add as many additional variables as you need to it**.

If you want to add factor variables, you can find a description of how to create drop-down lists here.

#### MISSING VALUES

**Missing values** should be kept as **blank cells**. The only exception is the SOURCE_AUTHOR for which unknown should be written.

#### OVERVIEW OF ALL PREDEFINED VARIABLES

The following **variables (columns)** are predefined in the data extraction templates. This overview can also be found in the Excel file (sheet "CODEBOOK"):

| **Variable name** | **Data type** | **Variable Description** |
|---|---|---|
| SOURCE_ID | Integer | Identification number of the input source |
| SOURCE_AUTHOR | String | First author of input source |
| SOURCE_YEAR | Integer | Year of publication |
| SOURCE_TITLE | String | Title of source |
| SOURCE_DOI | String | DOI of source (if available) |
| SOURCE_URL | String | URL of source, other than DOI (if available) |
| OPT_ACCESS_DATE | Date | Date the reviewer accessed the source, for websites & grey literature (format DD.MM.YYYY) |
| OPT_STUDY_TYPE | Factor | Study type of the input source |
| OPT_OTHER_STUDY_TYPE | String | Details if OPT_STUDY_TYPE = "Other" |
| REF_YEAR_START | Integer | Starting year of the data derived from the input source |
| REF_YEAR_END | Integer | End year of the data derived from the input source |
| REF_LOC_LEVEL | Factor | Location level investigated by the input source |
| REF_LOCATION | Factor | Geographic location for which the input source was used |
| REF_LOCATION_ISO3 | Factor | ISO3 code for country specified under REF_LOCATION (filled automatically) |
| REF_SEX | Factor | Sex of the population of the data derived from the input source |
| REF_AGE_START | Integer | Numerical value of starting age of the population of the data derived from the input source |
| REF_AGE_END | Integer | Numerical value of ending age of the population of the data derived from the input source |
| REF_NOTES | String | Additional remarks about input source |
| OPT_MEAN_AGE | Numerical | Mean age of study population. |
| OPT_MEDIAN_AGE | Numerical | Median age of study population. |
| OPT_SUBPOP | String | Sub-population that was investigated (e.g. pregnant, immuno-compromised) |
| OPT_CASES | Factor | Case definition (e.g. confirmed, probable, susceptible) |
| OPT_DISEASE | String | Disease details, values depend on hazard (e.g. invasive vs. non-invasive) |
| OPT_SEROTYPE | String | Serotype(s) of the pathogen reported in the source study |
| REF_SAMPLE_SIZE | Integer | Sample size of data derived from input source |
| VALUE_SYMPTOM | String | Symptom for which the input is given in this row |
| VALUE_UNIT | Factor | Unit the mean/ median duration refers to days, weeks, months or years |
| VALUE_X | Integer | Number of events in data derived from input source |
| VALUE_MEAN | Numerical | Mean estimate of the data derived from the input source |
| VALUE_MEDIAN | Numerical | Median estimate of the data derived from the input source |
| VALUE_DENOM | Integer | Population denominator for the value mean/median (e.g. 10,000 or 100,0000) |

| | | |
|---|---|---|
| VALUE_SE | Numerical | Standard error of the data derived from the input source |
| VALUE_P000 | Numerical | 0th percentile (minimum) of the data derived from the input source |
| VALUE_P2_5 | Numerical | 2.5th percentile of the data derived from the input source (lower limit 95%CI) |
| VALUE_P5 | Numerical | 5th percentile of the data derived from the input source |
| VALUE_P10 | Numerical | 10th percentile of the data derived from the input source |
| VALUE_P25 | Numerical | 25th percentile of the data derived from the input source |
| VALUE_P75 | Numerical | 75th percentile of the data derived from the input source |
| VALUE_P90 | Numerical | 90th percentile of the data derived from the input source |
| VALUE_P95 | Numerical | 95th percentile of the data derived from the input source |
| VALUE_P97_5 | Numerical | 97.5th percentile of the data derived from the input source (upper limit 95%CI) |
| VALUE_P100 | Numerical | 100th percentile (maximum) of the data derived from the input source |

Color coding:

| | |
|---|---|
| **Mandatory**, same for all rows from the same input source | **Optional**, same for all rows from the same input source |
| **Mandatory**, can differ per row from the same input source | **Optional**, can differ per row from the same input source |
| **At least 1 of them mandatory**, different for every row from the same input source | |
| **Mandatory**, different for every row from the same input source | **Optional**, different for every row from the same input source |

In the following, each of the variables is described in more detail.

#### DETAILS SOURCE VARIABLES

##### SOURCE_ID

The reviewers assign a **unique number** to each of the input sources. The same number is assigned to all rows concerning the same input source across all sheets, Example:

| SOURCE_ID | SOURCE_AUTHOR | ... | REF_AGE_START | REF_AGE_END | REF_SEX | REF_SAMPLE_SIZE | VALUE_X | ... |
|---|---|---|---|---|---|---|---|---|
| 1 | Smith, A | | 10 | 90 | All sexes | 100 | 20 | |
| 1 | Smith, A | | 10 | 90 | Male | 70 | 15 | |
| 1 | Smith, A | | 10 | 90 | Female | 30 | 5 | |
| 2 | Huber, B | | 20 | 39 | All sexes | 400 | 50 | |
| 2 | Huber, B | | 40 | 59 | All sexes | 600 | 100 | |

##### SOURCE_AUTHOR

Here the **name of the author** is specified in the following format:

- Firstname Lastname → "Lastname, F"
- Firstname Middlename Lastname → "Lastname, FM"

If there is no identifiable first author:

- Use the organization as author, format: Example Organization → "Example Organization, EO"
- If also no organization can be specified, use unknown

#### SOURCE_YEAR

Here the **year the input source was published** is specified. If not known: leave blank.

#### SOURCE_TITLE

Here the **English title of the source** is specified. If no English title is available, use the original language title and place it in square brackets [].

#### SOURCE_DOI

If available, copy the **DOI** of the input source here. If not, leave empty.

#### SOURCE_URL

If there is **another URL** besides the DOI available, copy it here. If not, leave empty.

### REFERENCE VARIABLES

#### REF_YEAR_START & REF_YEAR_END

Specify here the **period** over which the population has been observed (start & end year of this period).

If this is not stated in the input source, leave the fields empty.

If the source reports the results separately for multiple time periods, add a separate column for each of them (also add a row for total if the source reports the results in total). Example:

| SOURCE_ID | SOURCE_AUTHOR | … | REF_YEAR_START | REF_YEAR_END | … | REF_SAMPLE_SIZE | VALUE_X | … |
|---|---|---|---|---|---|---|---|---|
| 1 | Smith, A | | 2000 | 2020 | | 1000 | 30 | |
| 1 | Smith, A | | 2000 | 2005 | | 200 | 5 | |
| 1 | Smith, A | | 2006 | 2010 | | 100 | 15 | |
| 1 | Smith, A | | 2011 | 2015 | | 300 | 10 | |
| 1 | Smith, A | | 2016 | 2020 | | 400 | 50 | |

#### REF_LOC_LEVEL

Indicate here **the location level** the study was conducted on. The options are:

| Location level | How to add details |
|---|---|
| *National* | REF_LOCATION = country<br>*Special case: multi-country/regional studies that report results by country:*<br>Treat as national studies with separate rows for each country. |
| *Sub-national* | REF_LOCATION= country + specify details in REF_NOTES |
| *Regional* | *For multi-country/regional studies that report results only aggregated over all countries.*<br>Separate row for each country. Since it is not clear, how many cases came from each |

| | |
|---|---|
| | country, all countries receive the same incidence rate → VALUE_X is the same for each row. REF_SAMPLE_SIZE is the sum of the sample sizes of all countries included in the study. Specify which countries are included in REF_NOTES. |
| *Global* | Same as Regional |

Example:

| SOURCE_ID | SOURCE_AUTHOR | ... | REF_NOTES | ... | REF_LOC_LEVEL | REF_LOCATION | ... | REF_SAMPLE_SIZE | VALUE_X | ... |
|---|---|---|---|---|---|---|---|---|---|---|
| 1 | Smith, A | | | | National | Denmark | | 5 800 000 | 30 | |
| 2 | Huber, B | | Location: Vienna | | Sub-national | Austria | | 1 800 000 | 5 | |
| 3 | Brown, C | | North America: USA, Canada | | Regional | United States of America | | *332M (Population USA) + 38M (Population Canada) =* 370 000 000 | 200 | |
| 3 | Brown, C | | North America: USA, Canada | | Regional | Canada | | *332M (Population USA) + 38M (Population Canada) =* 370 000 000 | 200 | |
| 4 | Bauer, D | | European Union | | National | Belgium | | 11 000 000 | 10 | |
| 4 | Bauer, D | | European Union | | National | Italy | | 59 000 000 | 50 | |
| 4 | Bauer, D | | European Union | | National | France | | 67 000 000 | 100 | |

#### REF_LOCATION

Specify here the **country under investigation** (see REF_LOC_LEVEL for more details).

#### REF_LOCATION_ISO3

Here the **iso3 code** of the country under investigation is specified. This field is filled in automatically, please do not change the formula.

#### REF_SEX

*Short description: Sex of the population of the data derived from the input source.*

Here the **sex of the source population** (not the cases!!) is specified. The following drop-down options may be chosen:

| ***Input value*** | ***When to use*** |
|---|---|
| *All sexes* | If the source population consists of men and women or if the sex of the source |
| *Male* | If the source population consists only of men |
| *Female* | If the source population consists only of women |

If the sex of the source population is not given, leave this field empty.

If the input source reports the results separately for multiple sex categories, add a separate column for each of them (also add a row for total if the source reports the results in total). Example:

| SOURCE_ID | SOURCE_AUTHOR | ... | REF_SEX | REF_AGE_START | REF_AGE_END | REF_SAMPLE_SIZE | VALUE_X | ... |
|---|---|---|---|---|---|---|---|---|
| 1 | Smith, A | | All sexes | 10 | 90 | 100 | 20 | |
| 1 | Smith, A | | Male | 10 | 90 | 70 | 15 | |
| 1 | Smith, A | | Female | 10 | 90 | 30 | 5 | |

#### REF_AGE_START & REF_AGE_END

Here the **age range of the source population** (not the age of the cases!!) is specified, entering the age of the youngest person under "REF_AGE_START" and the age of the oldest person under "REF_AGE_END".

If this is not stated in the input source, leave the fields empty.

If the source study reports the results separately for multiple different age categories, add a separate column for each of them (also add a row for total if the source reports the results in total).

If age categories without any upper or lower bound are reported (i.e. ≤ 65 and >65), add in the appropriate column 0 or 125.

Example:

| SOURCE_ID | SOURCE_AUTHOR | ... | REF_SEX | REF_AGE_START | REF_AGE_END | REF_SAMPLE_SIZE | VALUE_X | ... |
|---|---|---|---|---|---|---|---|---|
| 1 | Smith, A | | All sexes | 19 | 100 | 1000 | 30 | |
| 1 | Smith, A | | All sexes | 19 | 29 | 200 | 5 | |
| 1 | Smith, A | | All sexes | 30 | 49 | 500 | 15 | |
| 1 | Smith, A | | All sexes | 50 | 69 | 300 | 10 | |
| 2 | Huber, B | | All sexes | 70 | 100 | 400 | 50 | |
| 2 | Huber, B | | All sexes | 40 | 59 | 600 | 100 | |
| 3 | Bauer, D | | All sexes | 0 | 65 | 600 | 100 | |
| 3 | Bauer, D | | All sexes | 66 | 125 | 600 | 100 | |

#### REF_SAMPLE_SIZE

The **sample size** always refers to the population from which the cases were identified. Examples:

- Population-based study: all persons living within the area in which cases were identified
- Outbreak reports: All persons that were exposed to the source of infection/ health hazard

If no sample size is given for population-based studies, the UN population estimates may be used instead: https://population.un.org/dataportal/data/indicators/49/locations/124/start/2000/end/2006/line/linetimeplot

!!! Keep in mind: At least one of the following MUST be specified:
REF_SAMPLE_SIZE **AND** VALUE_X **OR** VALUE_MEAN **OR** VALUE_MEDIAN !!!

#### REF_NOTES

Here any relevant additional remarks about the study, such as further details about the study location, may be specified.

### VALUE VARIABLES

#### VALUE_SYMPTOM

*(Only relevant for duration & sequelae)*

A separate row needs to be filled out for **each symptom** the study describes. Here, the symptom for which the data is entered in the respective row is specified.

Example:

| SOURCE_ID | SOURCE_AUTHOR | … | REF_SAMPLE_SIZE | VALUE_SYMPTOM | VALUE_X | … |
|---|---|---|---|---|---|---|
| 1 | Smith, A | | 100 | Fever | 20 | |
| 1 | Smith, A | | 100 | Diarrhea | 15 | |
| 1 | Smith, A | | 100 | Fatigue | 50 | |

#### VALUE_UNIT

*(Only relevant for duration)*

Factor variable, specify here which of the following **time units** the mean/median duration refers to:

- Days
- Weeks
- Months
- Years
- Other → Elaborate under REF_NOTES

Leave empty if not known.

Example:

| SOURCE_ID | SOURCE_AUTHOR | … | REF_SAMPLE_SIZE | VALUE_SYMPTOM | VALUE_X | … |
|---|---|---|---|---|---|---|
| 1 | Smith, A | | 100 | Fever | 20 | |
| 1 | Smith, A | | 100 | Diarrhea | 15 | |
| 1 | Smith, A | | 100 | Fatigue | 50 | |

#### VALUE_X

*(Only relevant for incidence, prevalence, mortality, sequelae, duration)*

Specify the **number of events of interest**, which are:

- Incidence & prevalence: number of cases
- Mortality: number of deaths
- Sequelae: number of persons experiencing the symptom
- Duration: duration of the symptom (in combination with the VALUE_UNIT variable).

!!! Keep in mind: At least one of the following MUST be specified:
REF_SAMPLE_SIZE **AND** VALUE_X **OR** VALUE_MEAN **OR** VALUE_MEDIAN !!!

#### VALUE_MEAN

*Short description: Mean estimate of the data derived from the input source*

!!! Keep in mind: At least one of the following MUST be specified:
REF_SAMPLE_SIZE **AND** VALUE_X **OR** VALUE_MEAN **OR** VALUE_MEDIAN !!!

#### VALUE_MEDIAN

*Short description: Median estimate of the data derived from the input source*

!!! Keep in mind: At least one of the following MUST be specified:
REF_SAMPLE_SIZE **AND** VALUE_X **OR** VALUE_MEAN **OR** VALUE_MEDIAN !!!

#### VALUE_DENOM

(Only relevant for incidence, prevalence & mortality)

Here the **population denominator** can be specified - only relevant if the input source reports a mean or median value. Not all sources will use the same population denominator for these values, some might e.g. report results per 10,000 and others per 100,000 persons. Please do not transform the values in any way, but report them exactly as they are specified by the input source and indicate the respective population denominator here.

If the mean or median refers to a percentage, the population denominator is 100.

#### VALUE_SE

Here the **standard error** of the outcome can be specified. Only needs to be filled out if stated in the input source.

#### VALUE_P0 → VALUE_P100

Here the **X$^{th}$ percentile** of the data derived from the input source can be specified. Only needs to be filled out if stated in the input source.

Reminder:

- VALUE_P2_5: lower 95% confidence interval limit
- VALUE_P97_5: upper 95% confidence interval limit

### OPTIONAL VARIABLES

#### OPT_ACCESS_DATE

Here the date the reviewer accessed the source may be filled out if the reviewer believes that the input source could be changed or removed in the future. This is relevant for, e.g. websites or grey literature.

#### OPT_STUDY_TYPE & OPT_OTHER_STUDY_TYPE

Specify here what **type of study** the input source is. The following drop-down options are given:

- Outbreak report
- Cross-sectional study
- Active surveillance
- Passive surveillance
- Cohort study
- Case series
- Modelling study
- Other → elaborate under "OPT_OTHER_STUDY_TYPE"

#### OPT_MEAN_AGE / OPT_MEDIAN_AGE

Here, the **mean or median age** of the source population (not the cases!!) may be stated. These variables can be used if the input source does not specify any age range.

#### OPT_SUBPOP

If the study did not concern the general population but a **specific sub-population**, e.g. immuno-compromised persons, this could be indicated here.

#### OPT_CASES

Here the **case definition** can be specified. The following drop-down values can be selected:

- All
- Confirmed
- Suspected
- Other → elaborate under REF_NOTES

If a study differentiates between e.g. probable and confirmed cases, a separate row for each case definition may be specified (also add a row for all cases if the source reports the results in total).

Example:

| SOURCE_ID | … | OPT_CASES | … | REF_SAMPLE_SIZE | VALUE_X | VALUE_MEAN | … |
|---|---|---|---|---|---|---|---|
| 1 | | All | | 50 | 10 | 0.2 | |
| 1 | | confirmed | | 50 | 3 | 0.06 | |
| 1 | | probable | | 50 | 7 | 0.14 | |

#### OPT_DISEASE

Depending on the hazard under investigation, more details about the disease type investigated may be informative and can be specified here; examples: invasive vs non-invasive; congenital vs non-congenital.

#### OPT_SEROTYPE

Depending on the hazard under investigation, the serotype(s) investigated in the study may be informative and can be specified here.
