## Supplementary material for "Computational framework for the World Health Organization estimates of the global, regional and national burden of foodborne diseases 2026 edition": S9 Appendix

### CTTF

### **Aflatoxin B1**
### Disease Model

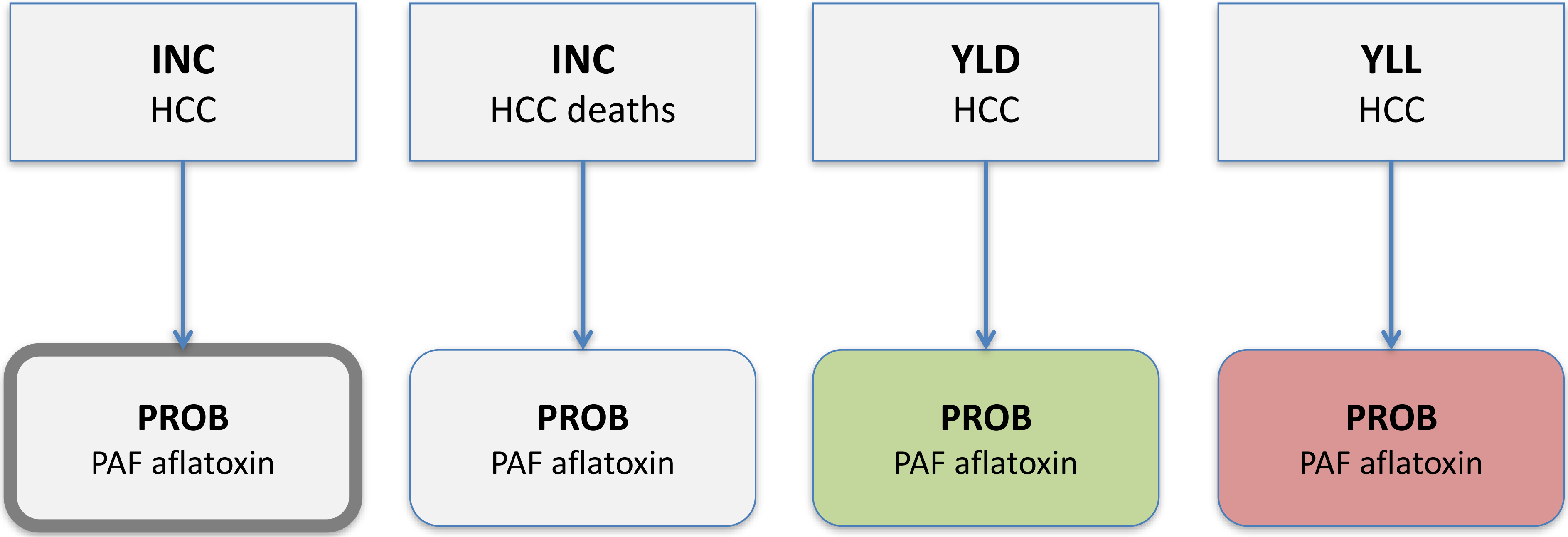

### **Aflatoxin M1**
### Disease Model

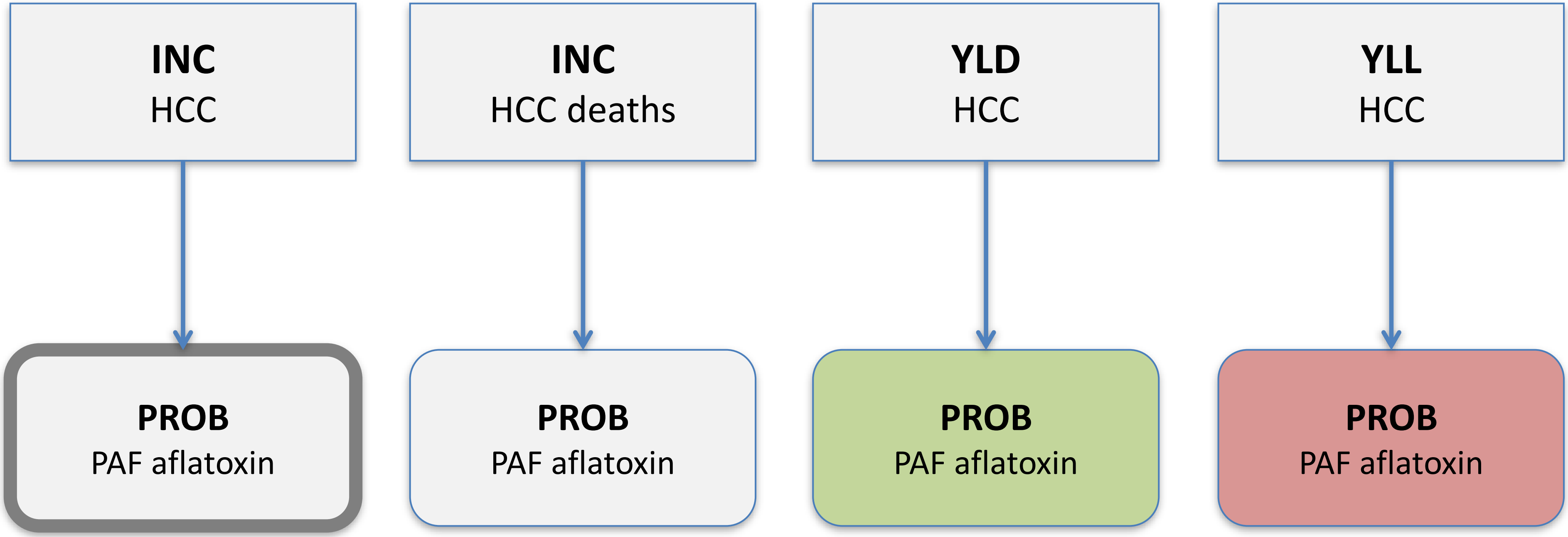

### Cyanide in cassava
Disease Model

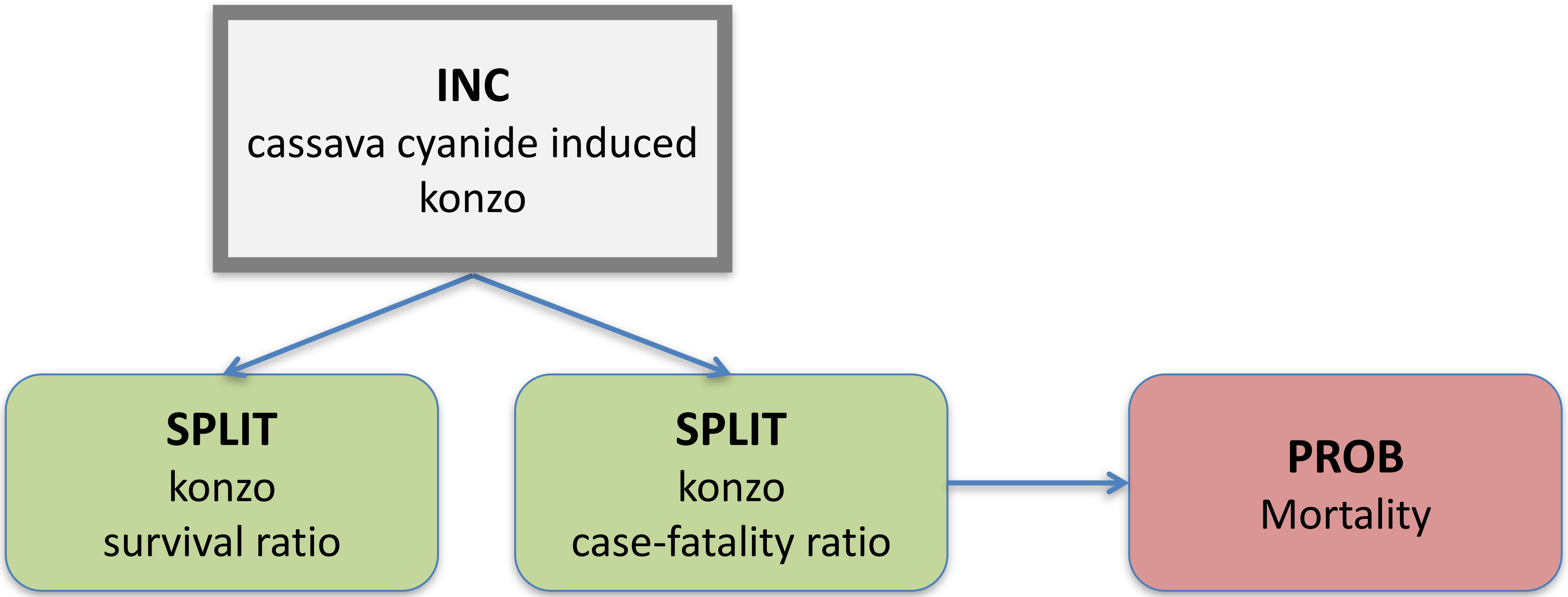

### **Dioxin**
### Disease Model

**INC**

Dioxin-induced impaired male fertility

↓

**PROB**

cases/100k births to cases/100k population correction factor

### Arsenic
### Disease Model

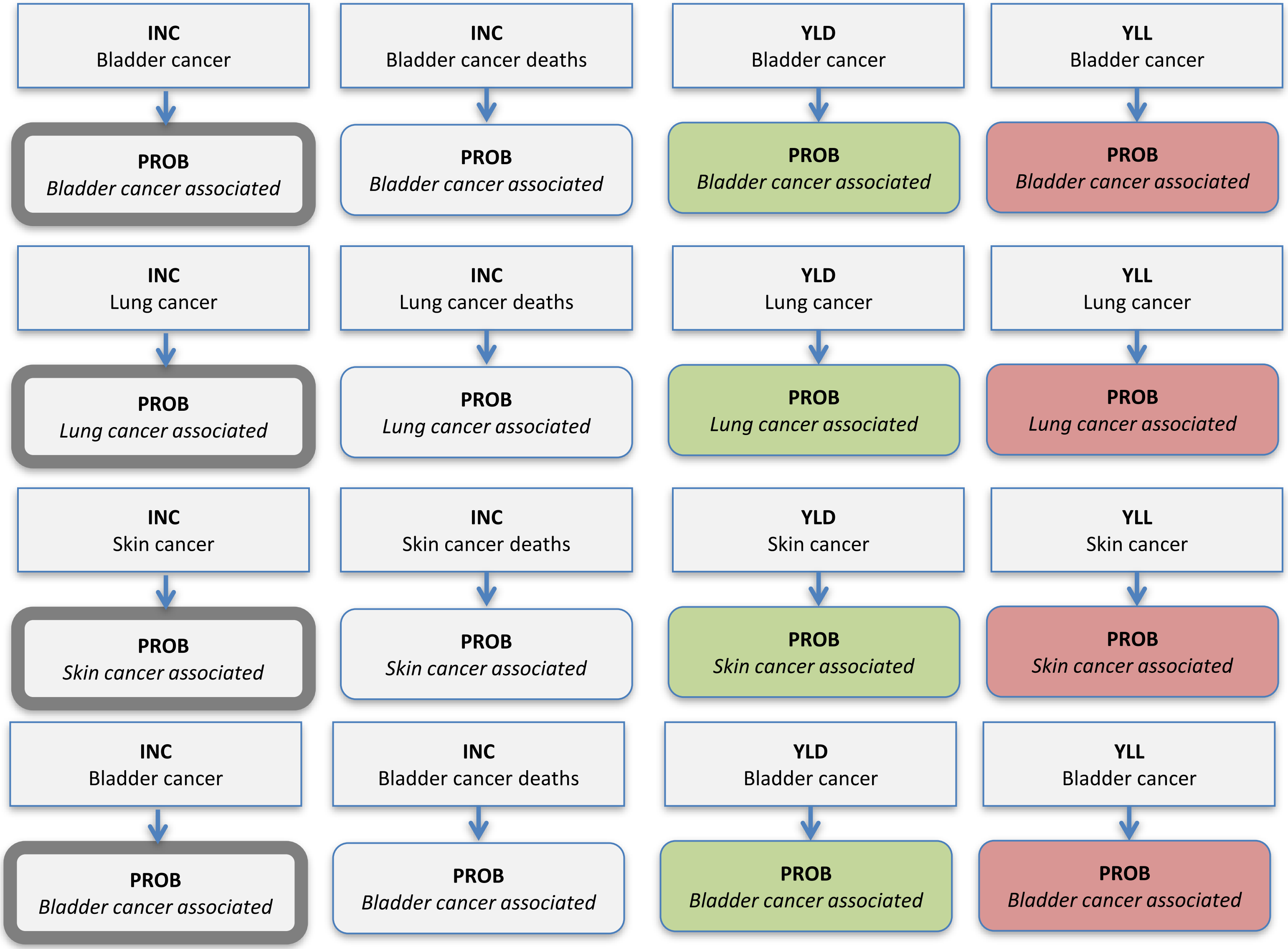

### **Cadmium**
### Disease Model

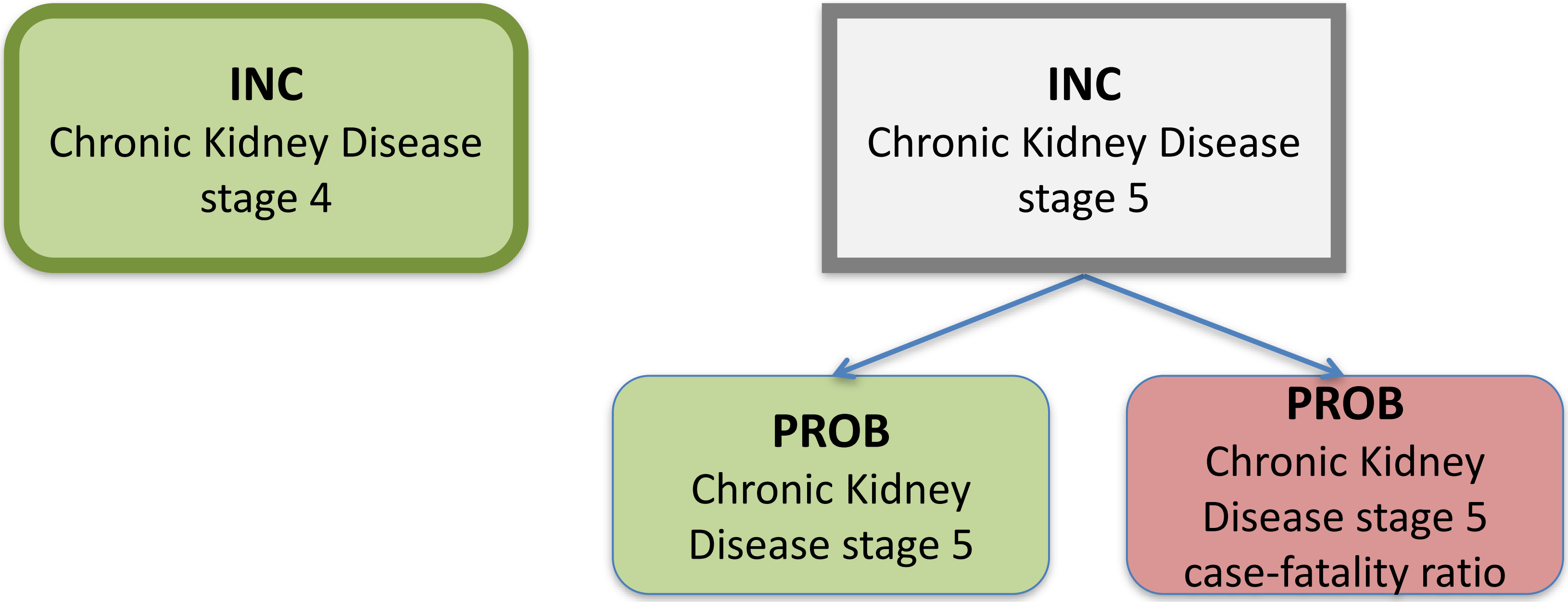

### **Lead**
### Disease Model

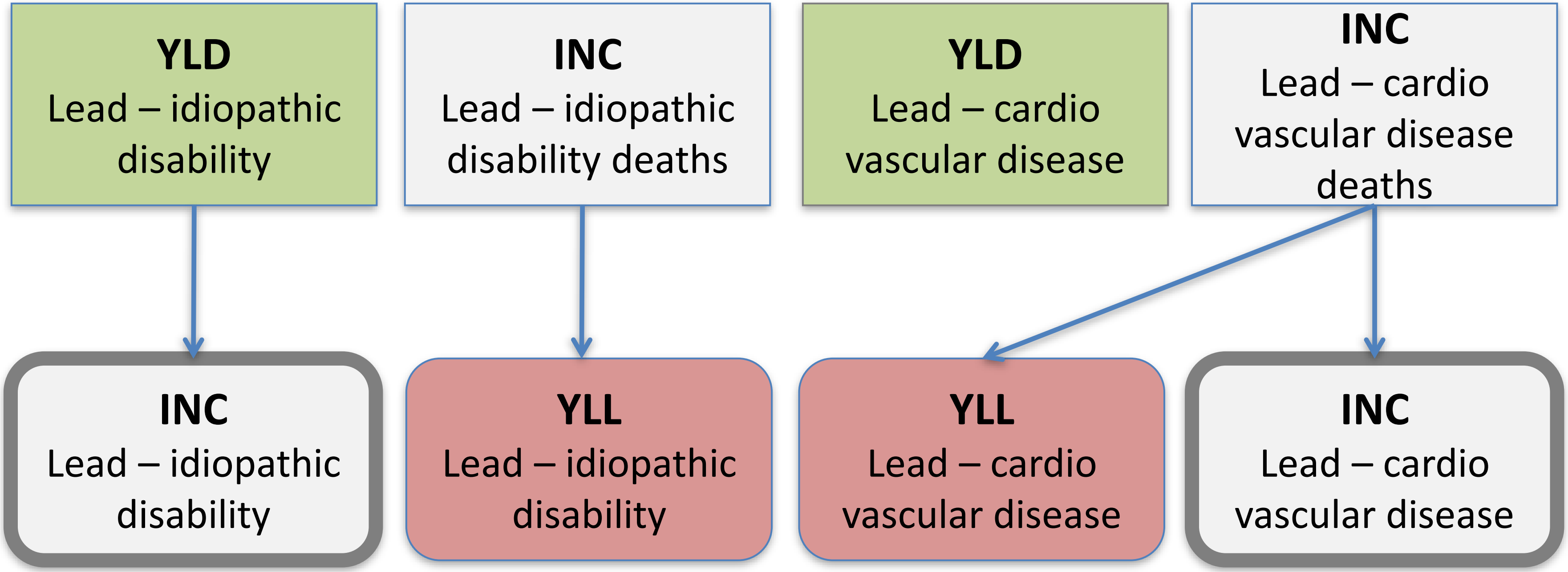

### **Methyl mercury**
### Disease Model

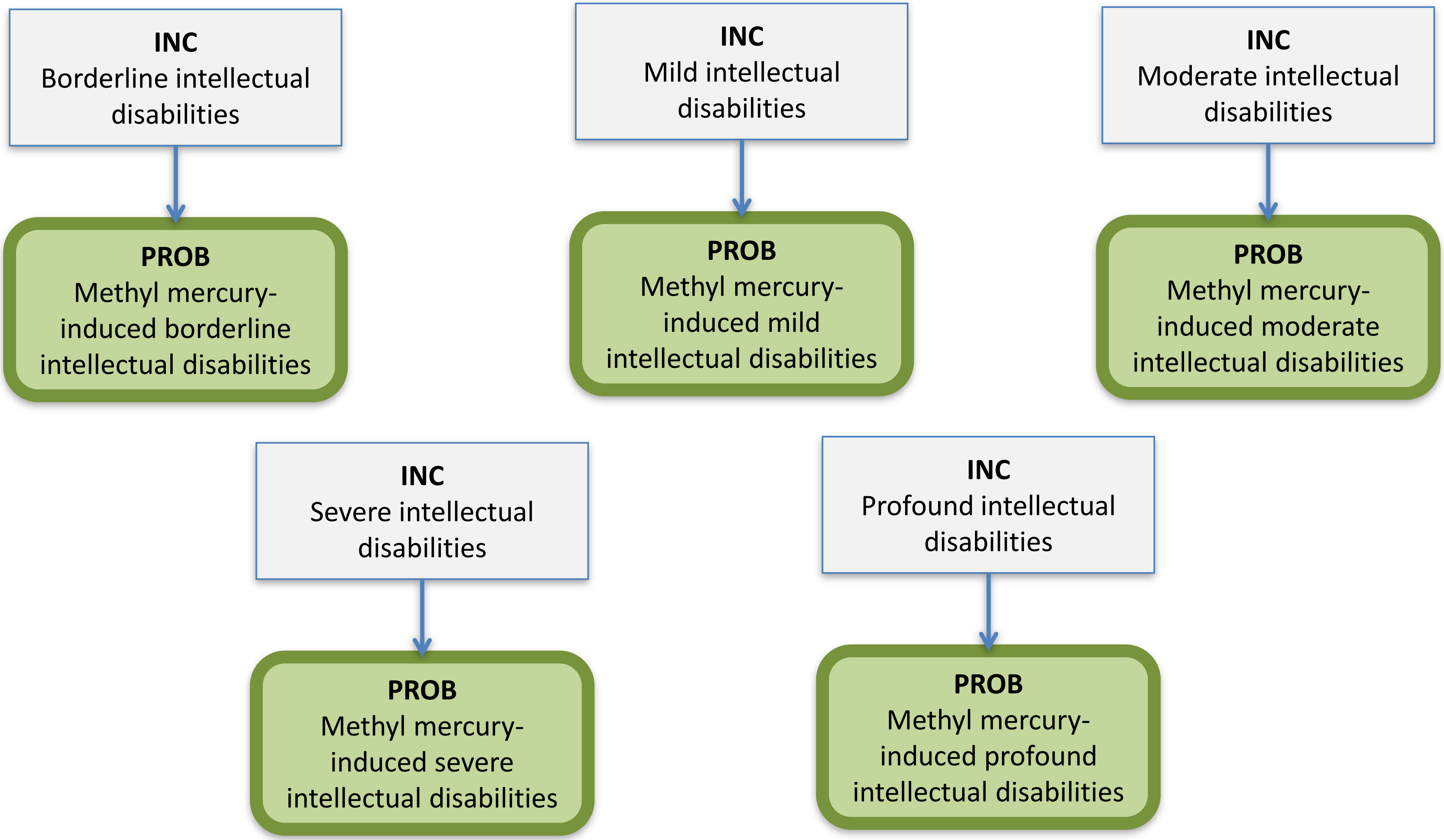

### **Peanut allergen**
### Disease Model

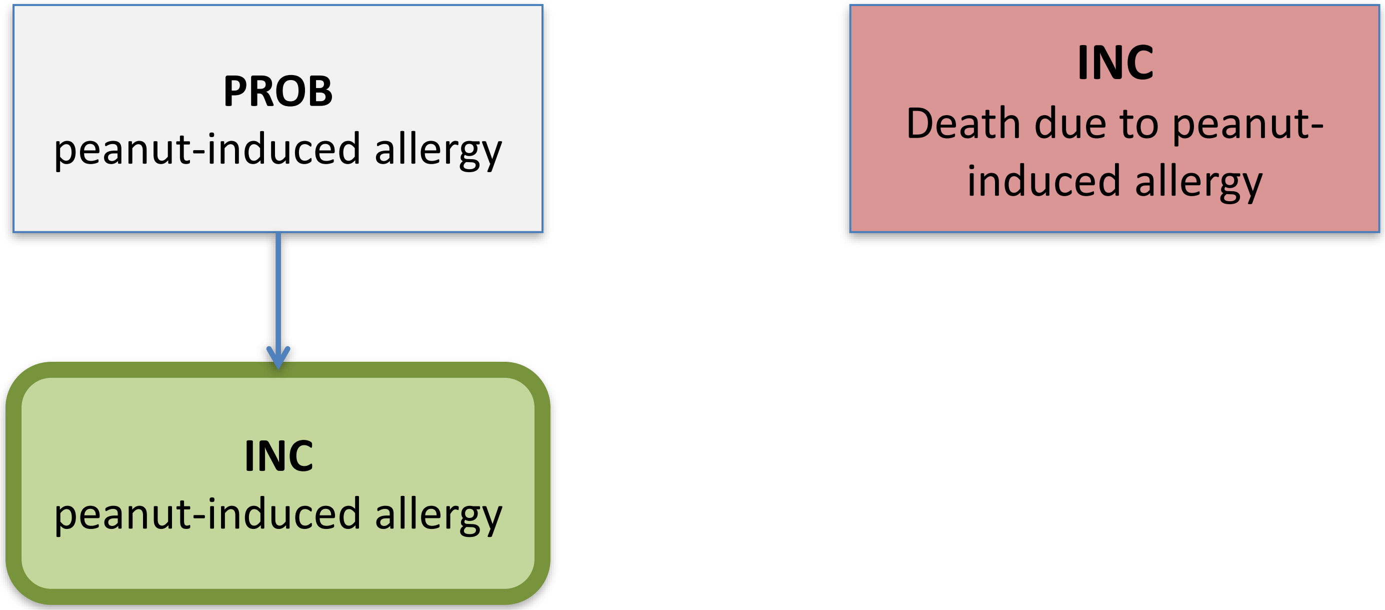

### EDTF

### ***Clostridium botulinum***
### Disease Model

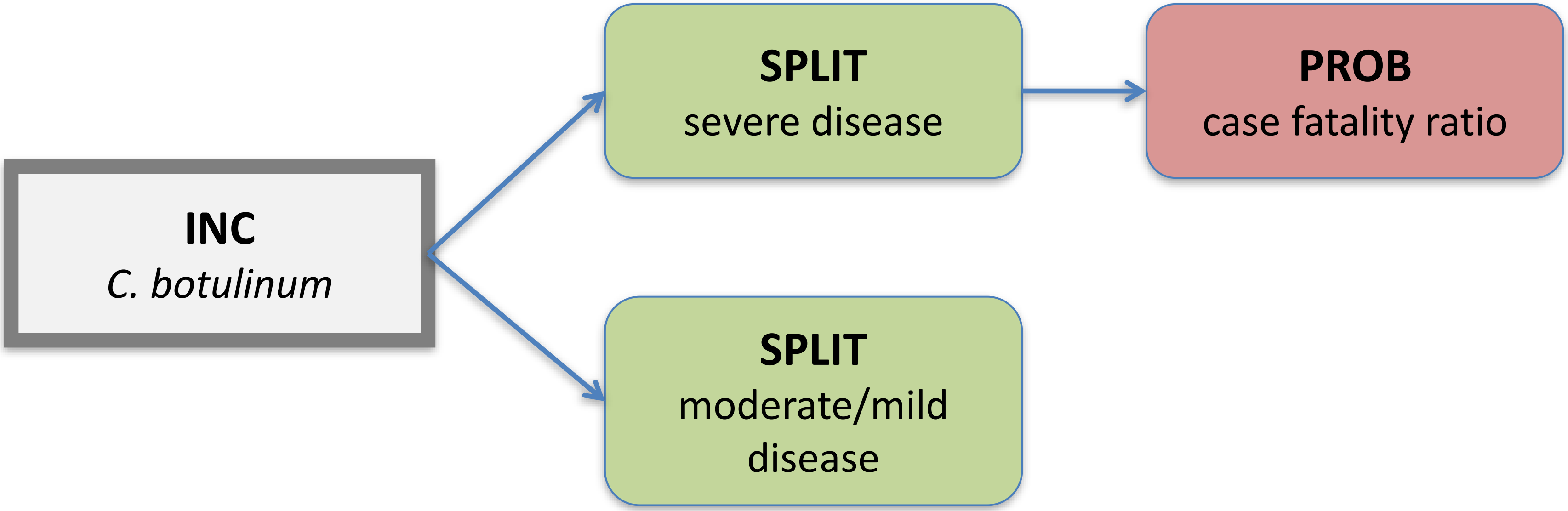

### ***Brucella* spp.**
### Disease Model

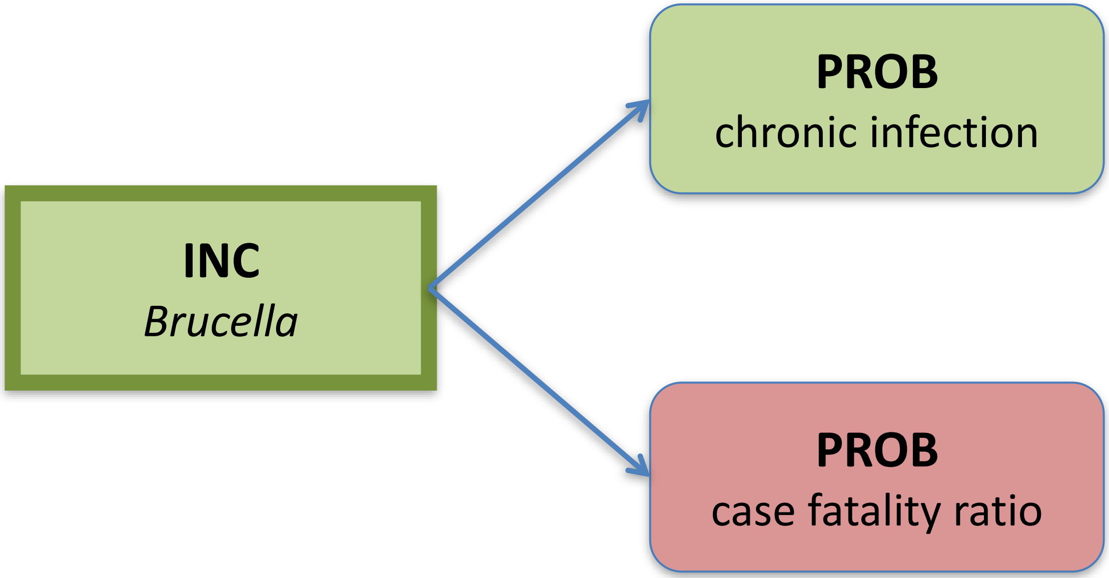

### *Campylobacter* spp.
### Disease Model

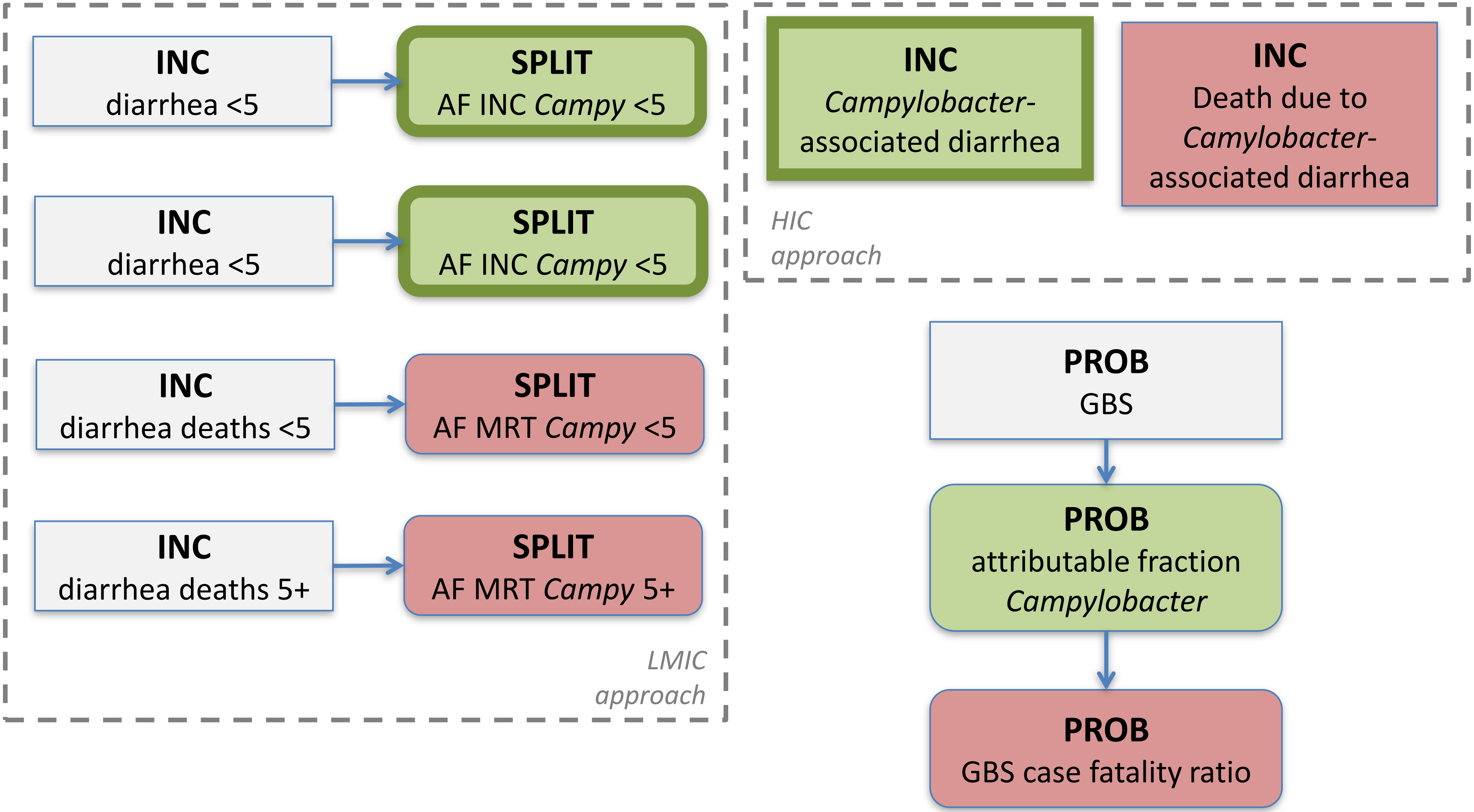

### *Cryptosporidium* spp.
### Disease Model

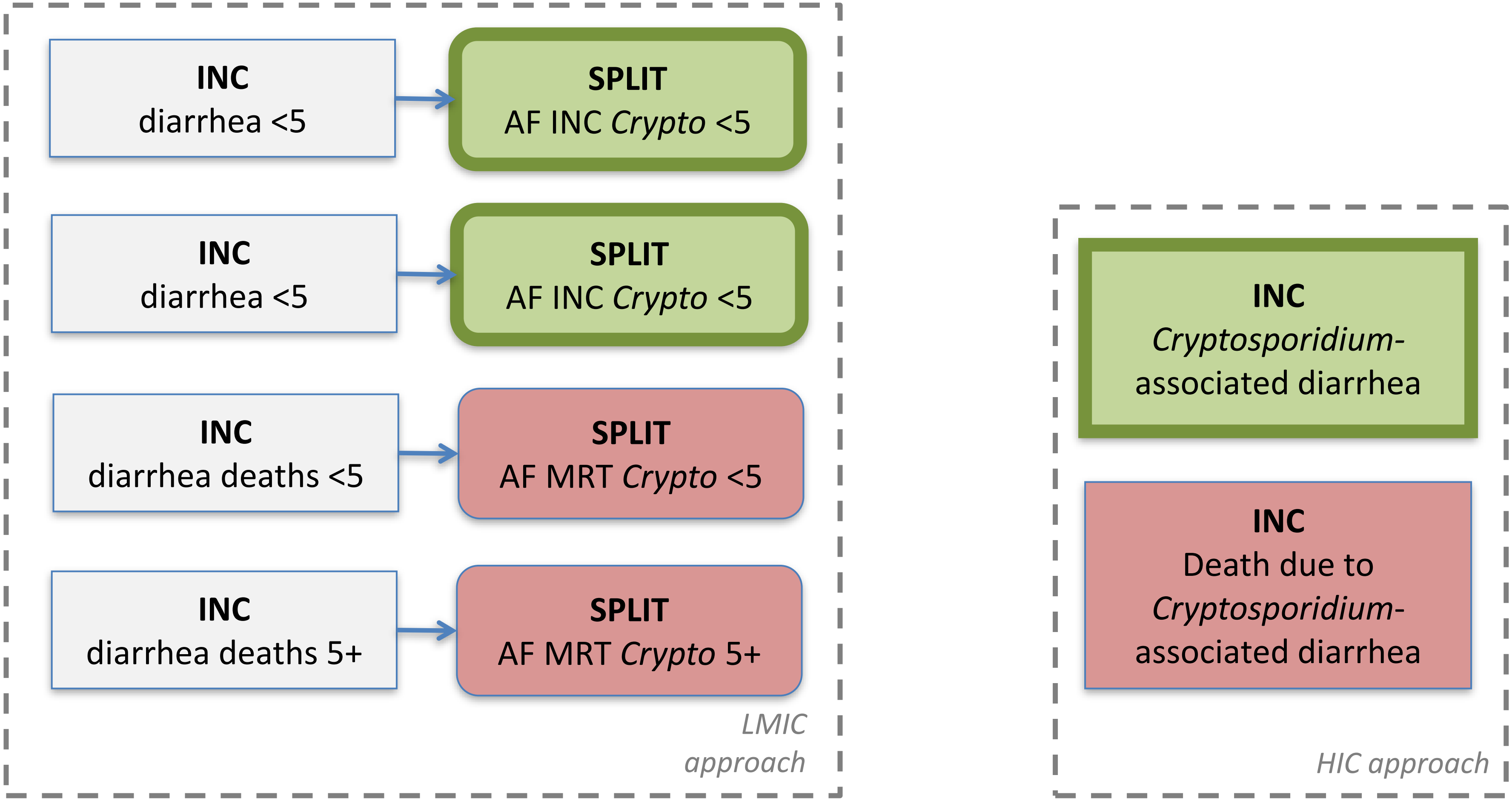

### *Cyclospora* spp. Disease Model

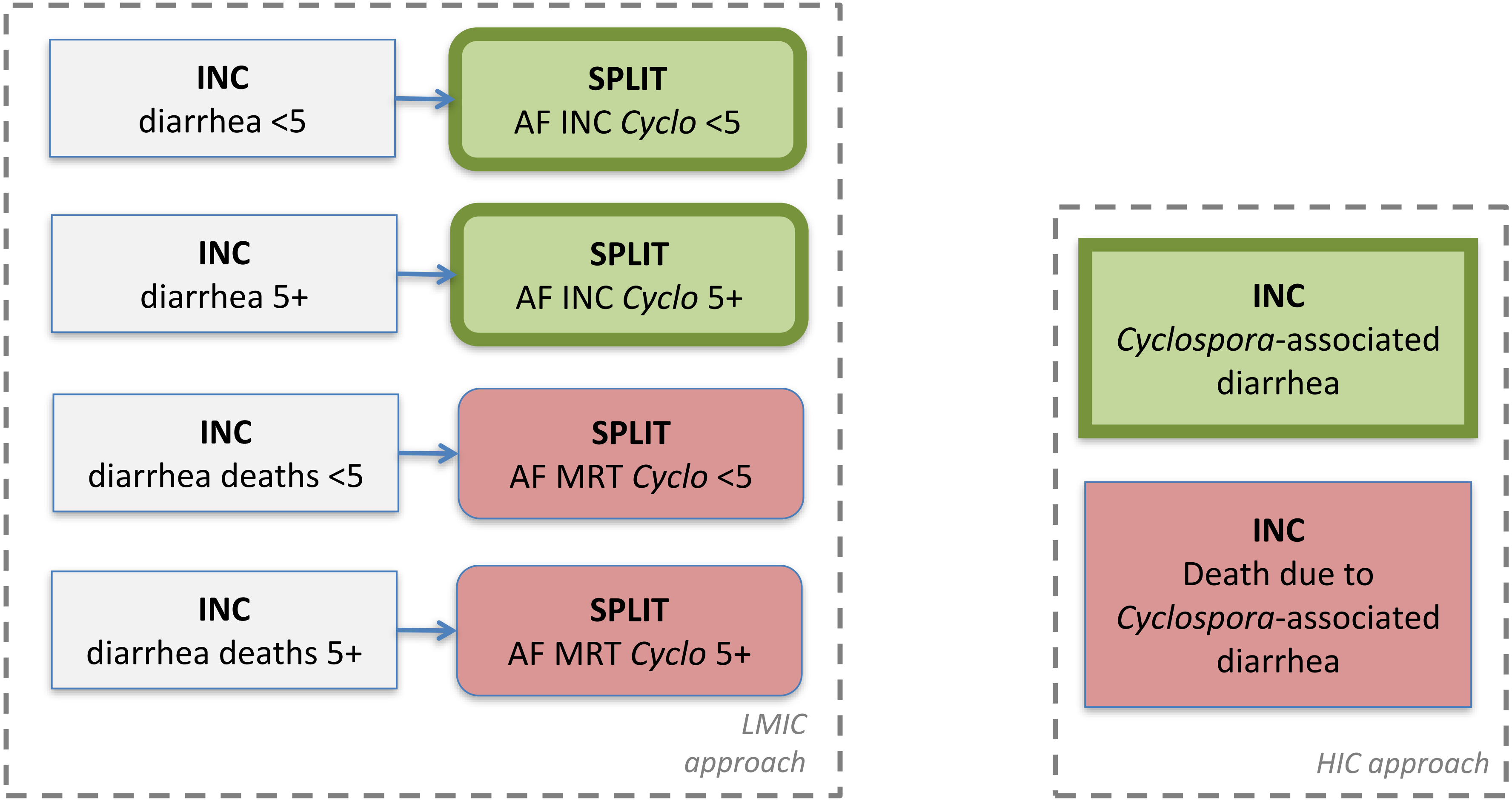

### **EAEC**
### Disease Model

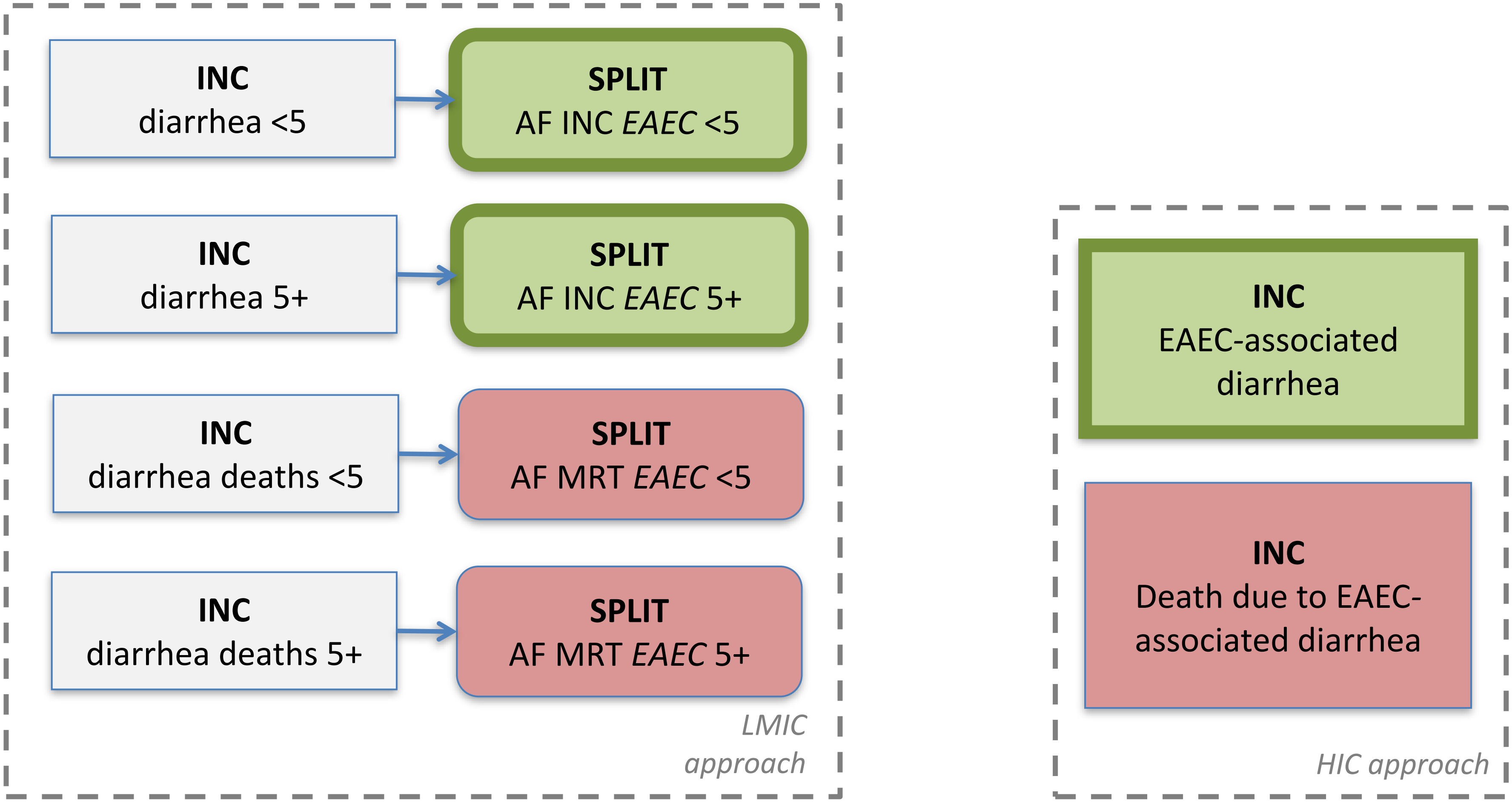

### *Entamoeba histolytica* Disease Model

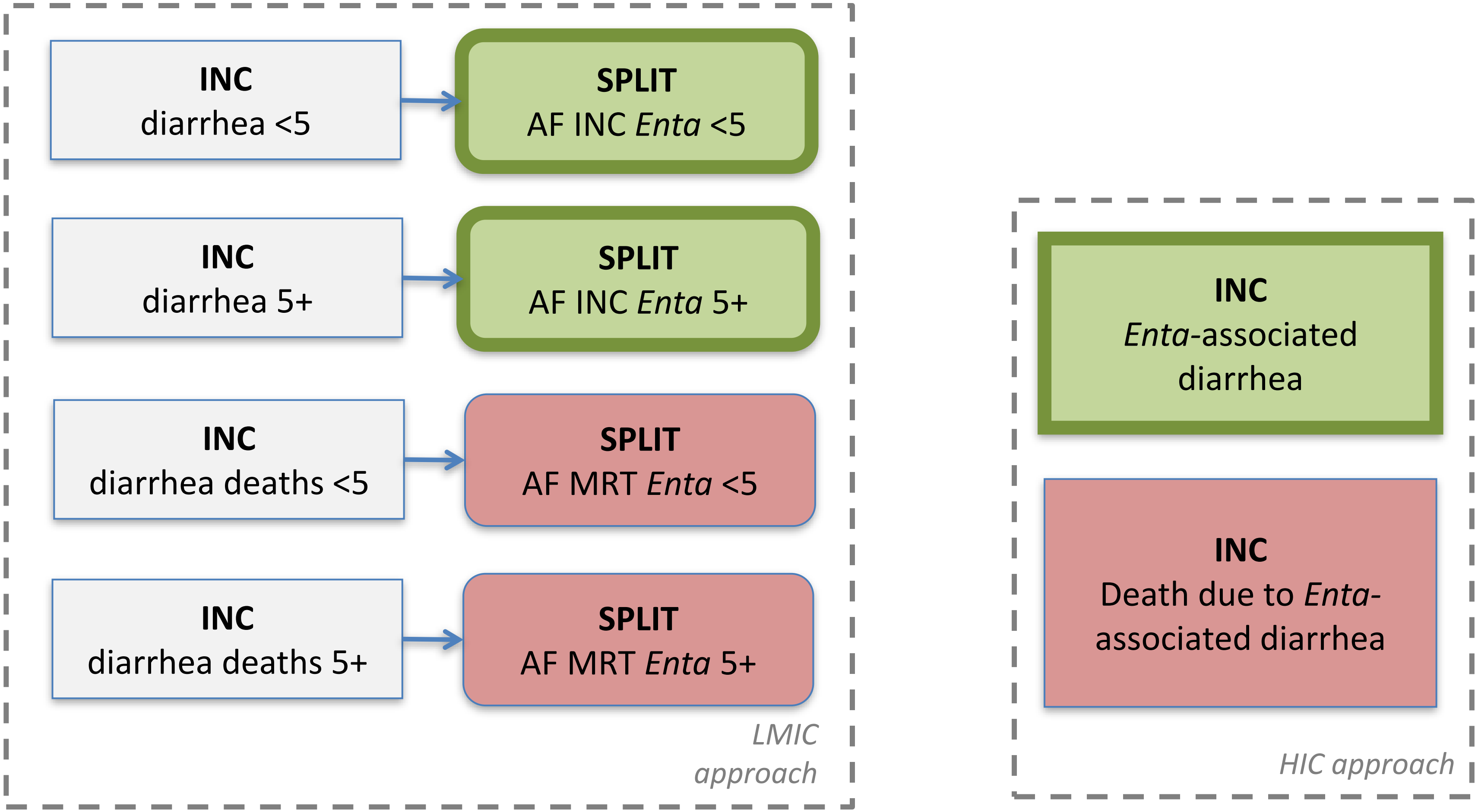

### EPEC
Disease Model

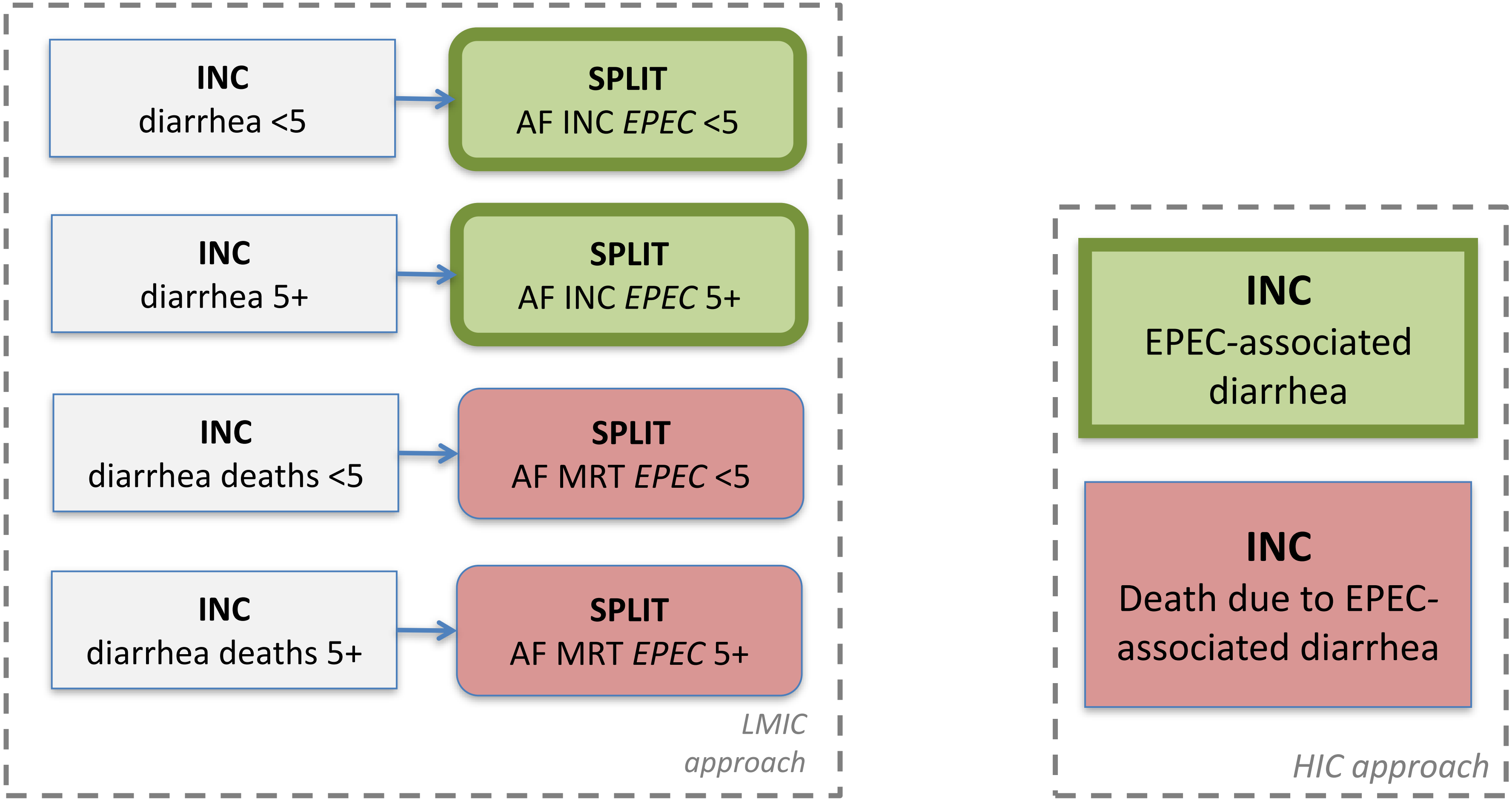

### **ETEC**
### Disease Model

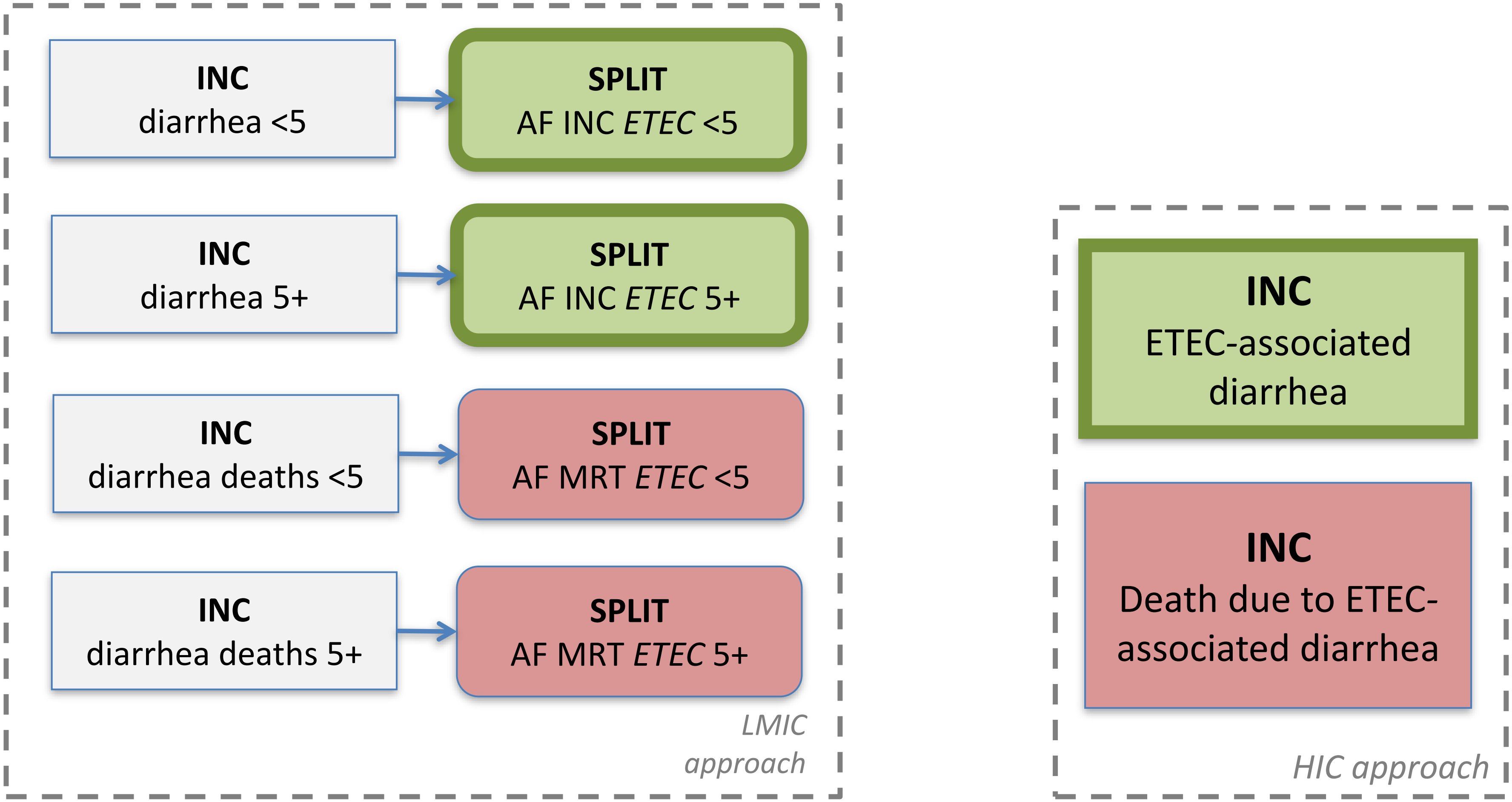

### *Giardia* spp.
### Disease Model

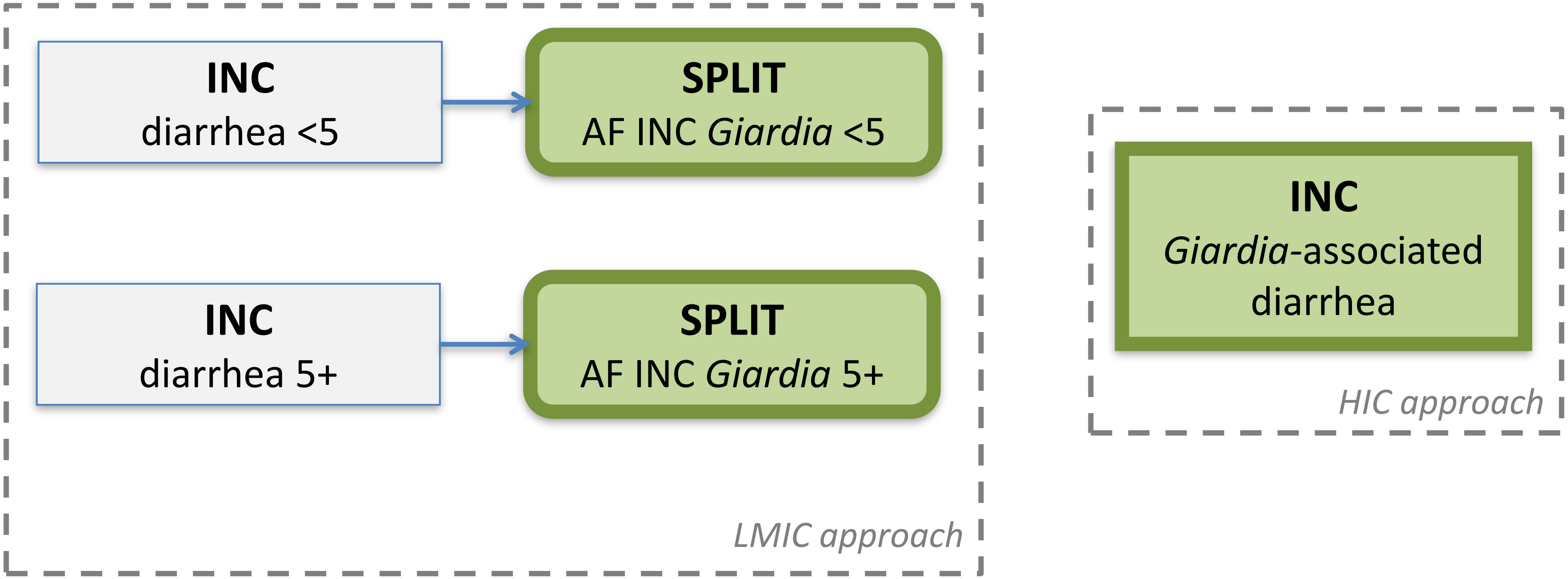

### Norovirus
### Disease Model

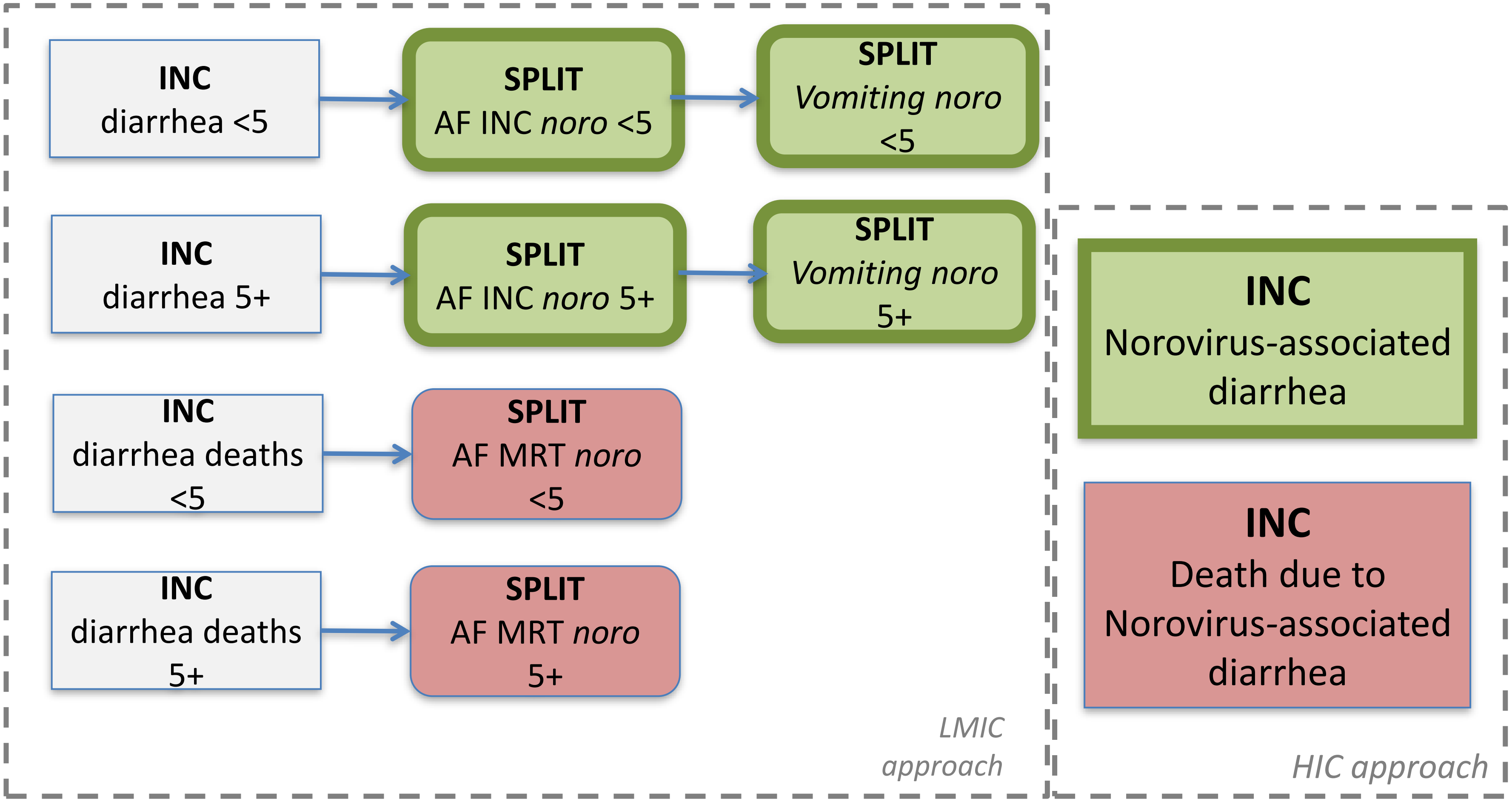

### **Rotavirus**
### Disease Model

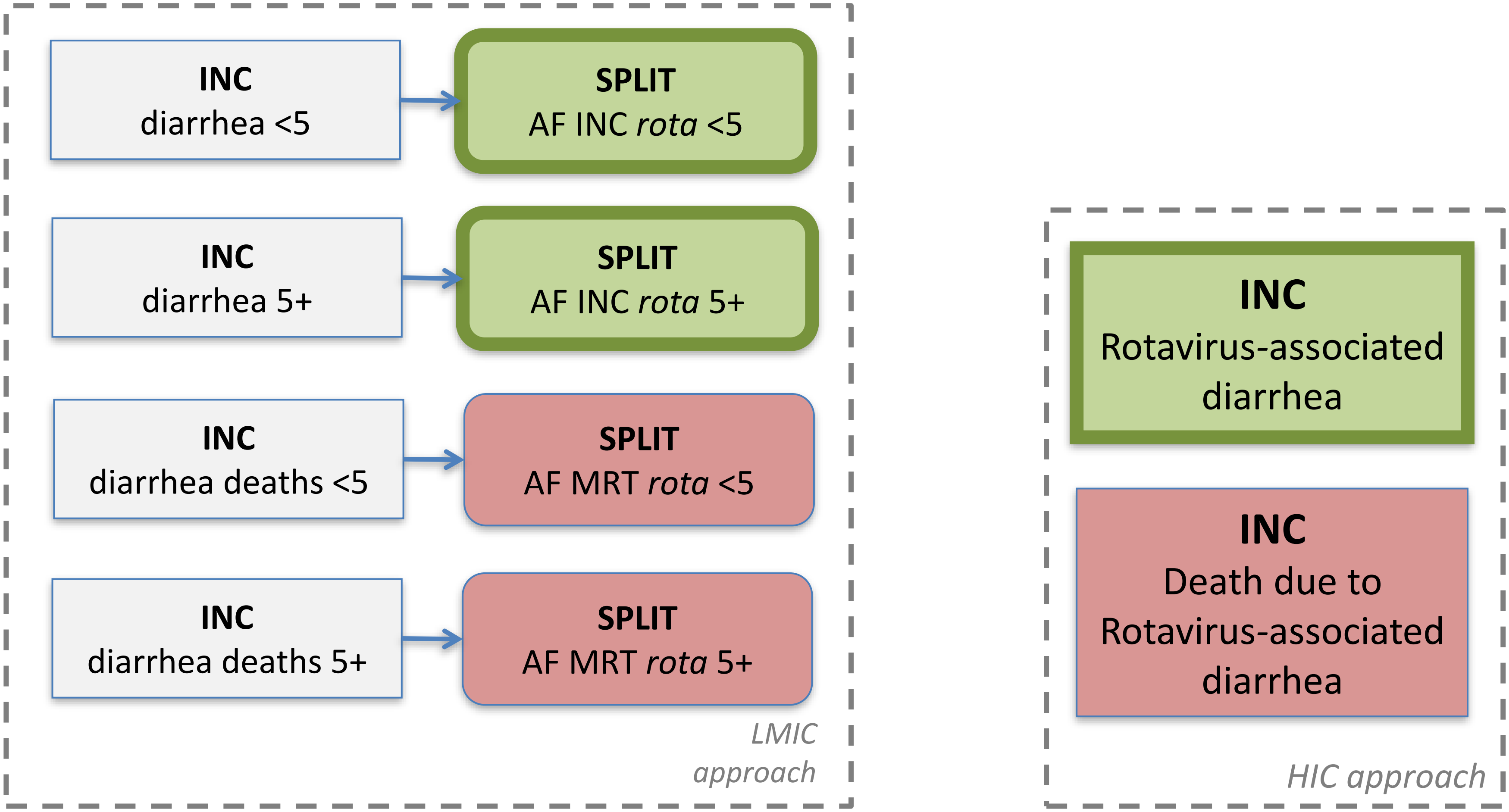

### *Salmonella enterica* Disease Model

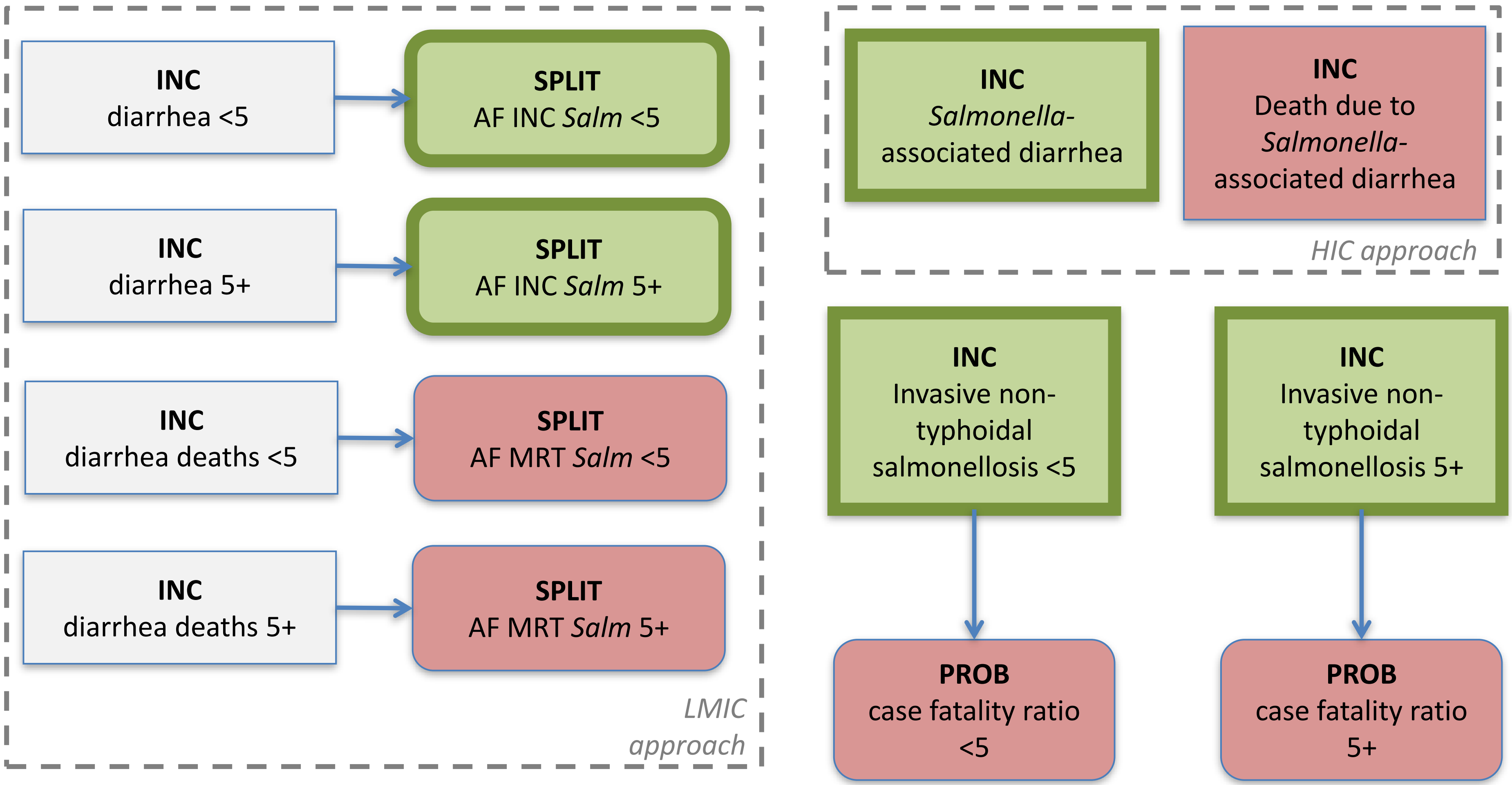

### *Shigella* spp.
### Disease Model

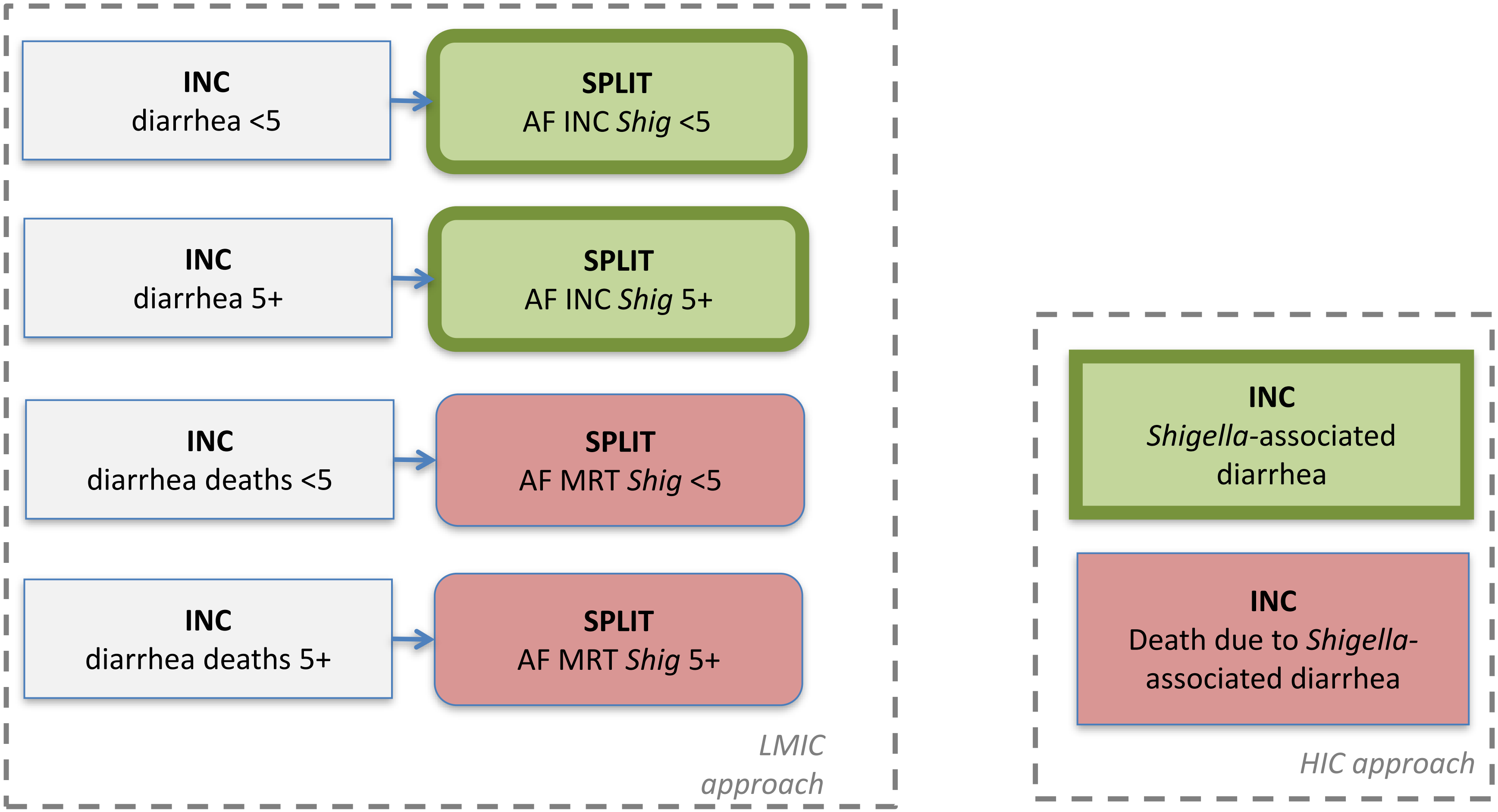

### STEC
### Disease Model

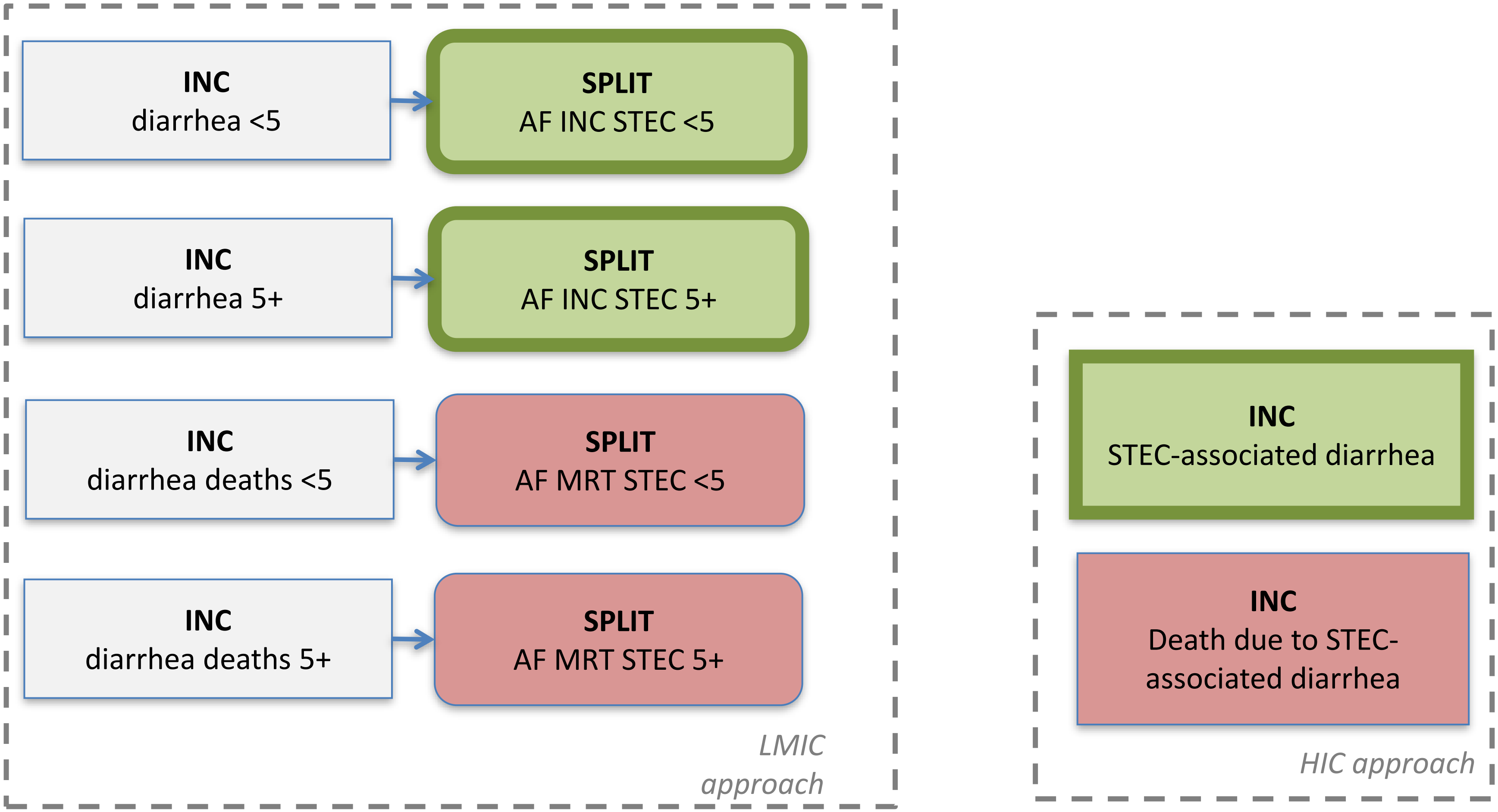

### **STEC (continued)** Disease Model

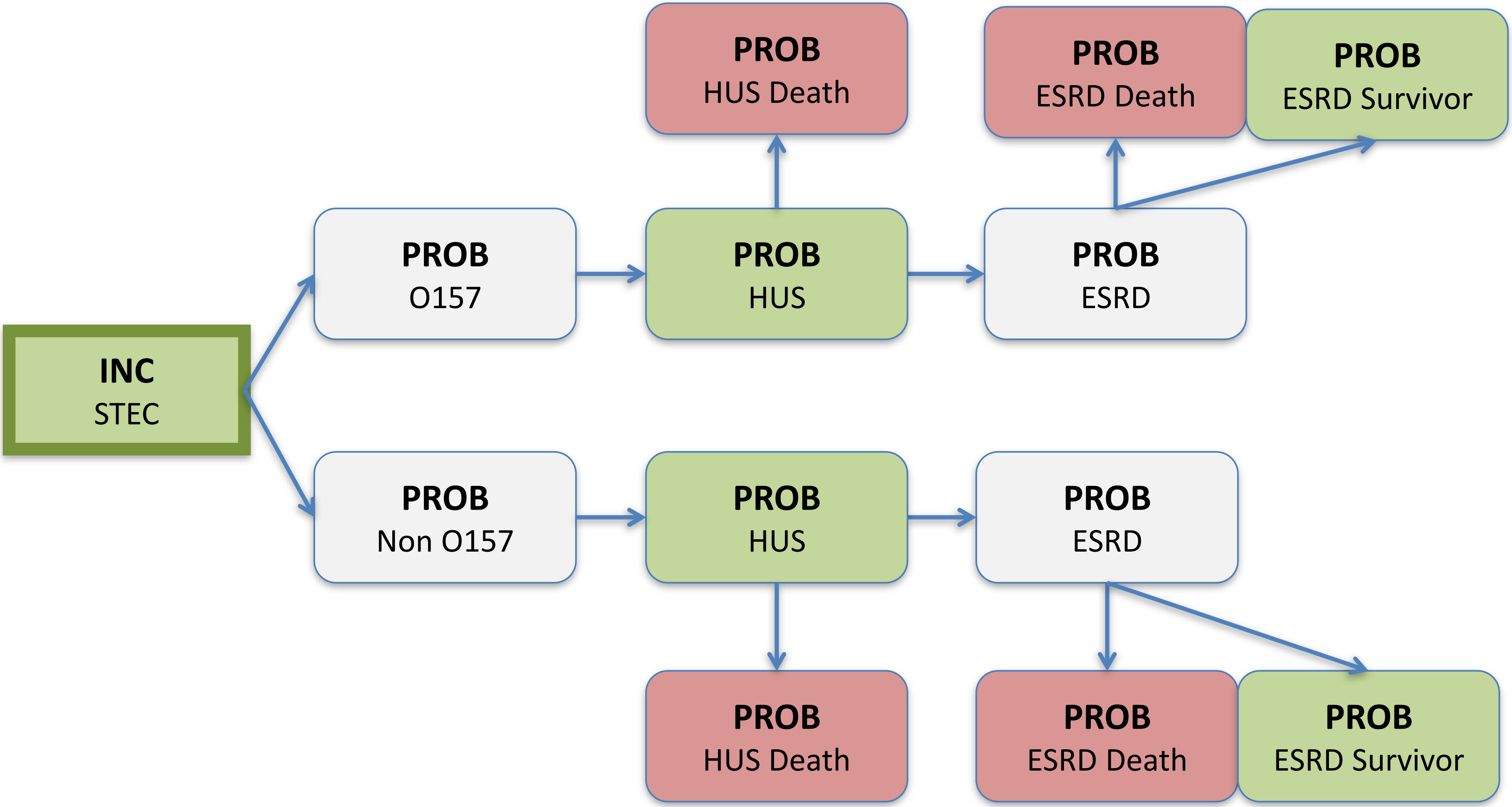

### *Vibrio cholerae*
### Disease Model

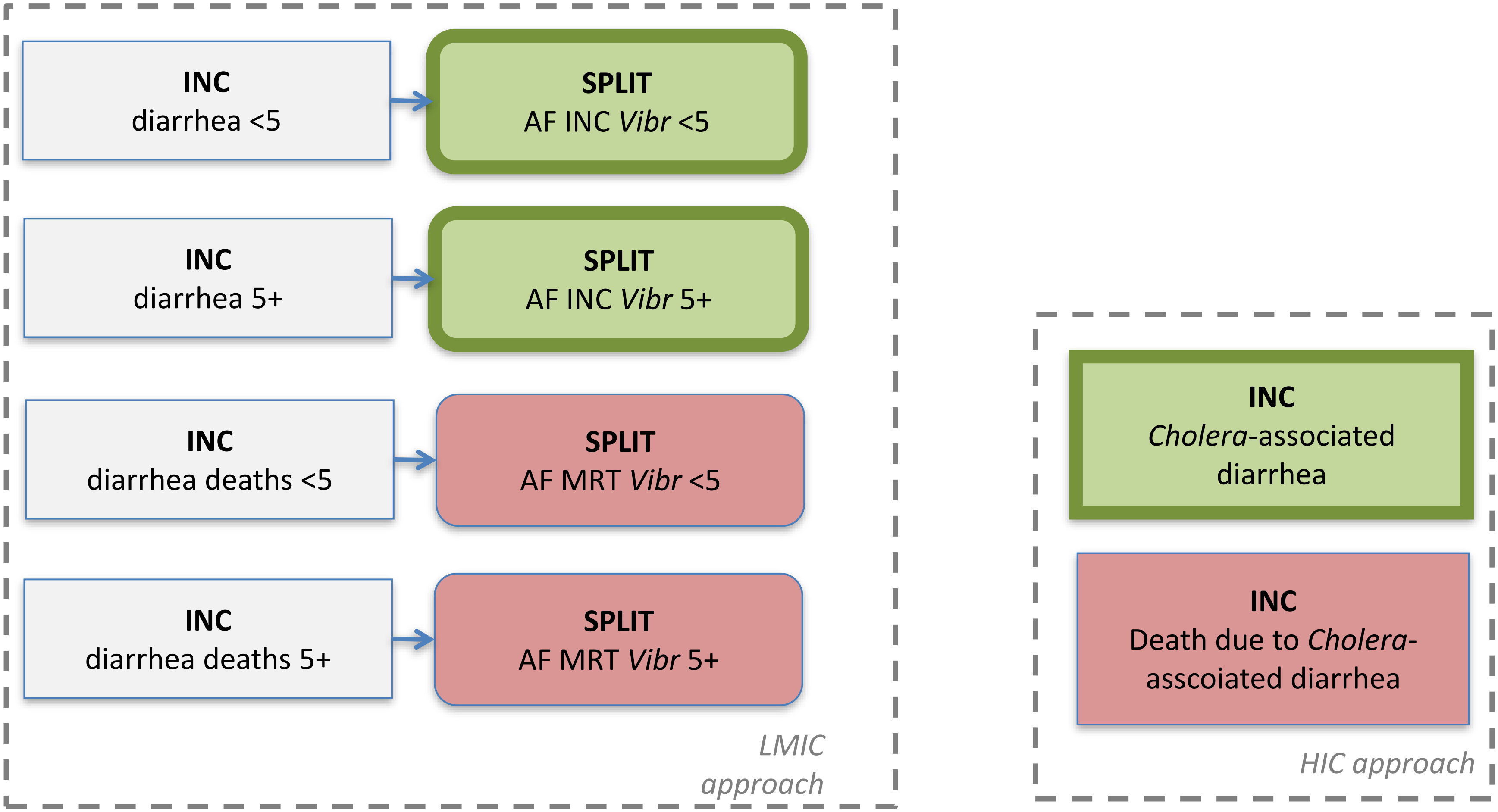

### **Hepatitis A virus**
### Disease Model

**INC**
Moderate hepatitis A

**INC**
Severe hepatitis A

**INC**
Hepatitis A
deaths

### *Listeria monocytogenes* Disease Model

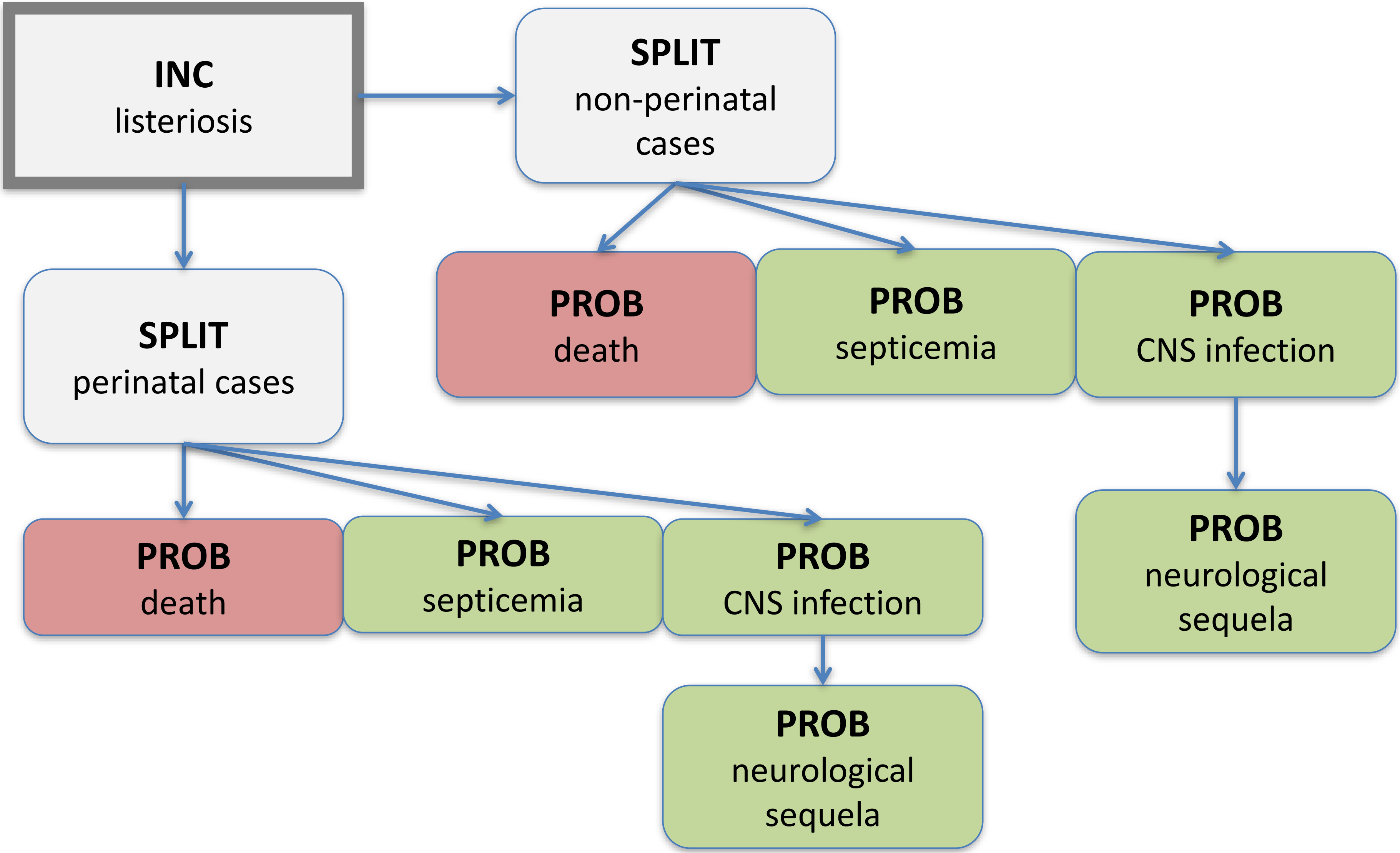

### *Salmonella* Typhi
### Disease Model

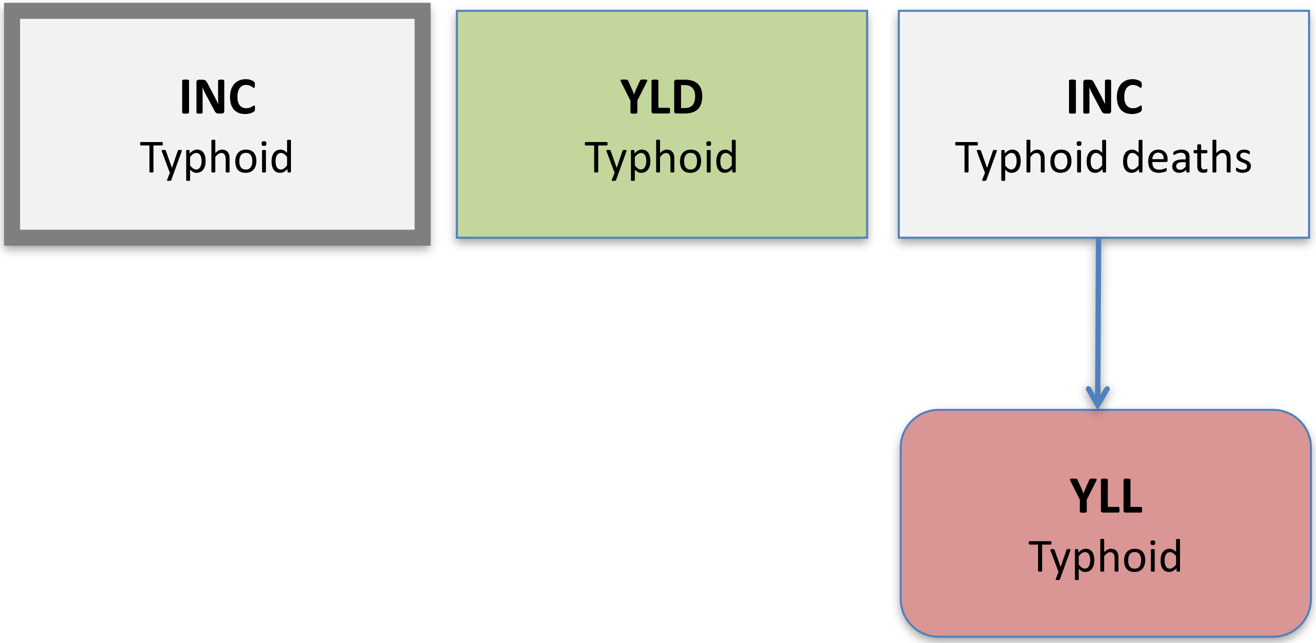

### *Salmonella* Paratyphi Disease Model

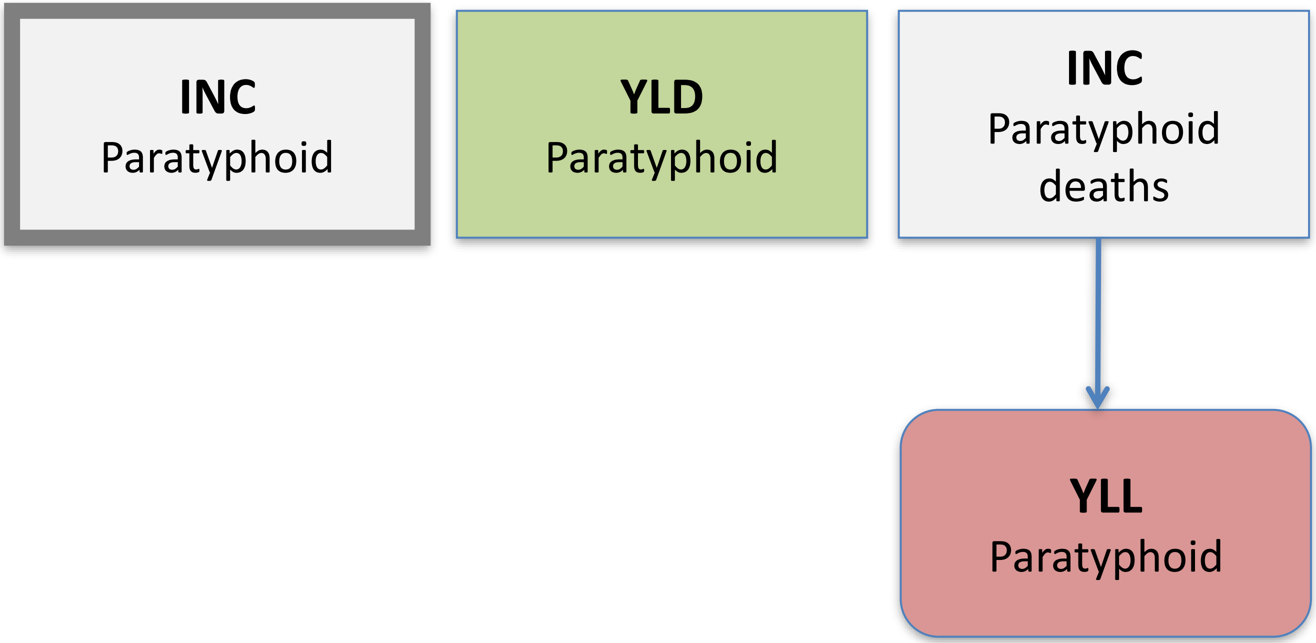

### *Mycobacterium bovis, caprae, and orygis* Disease Model

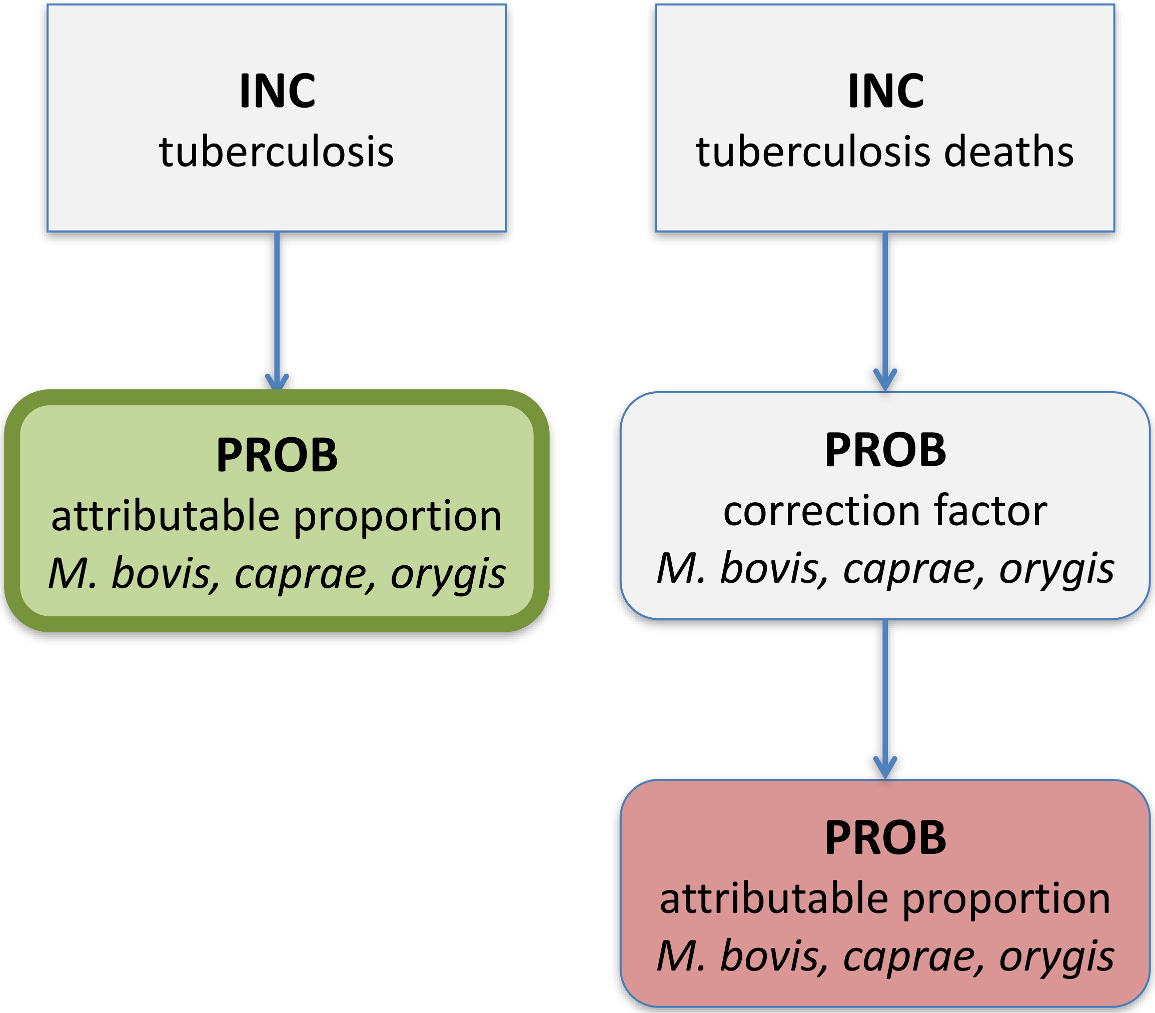

### PDTF

### *Ascaris spp.* Disease Model

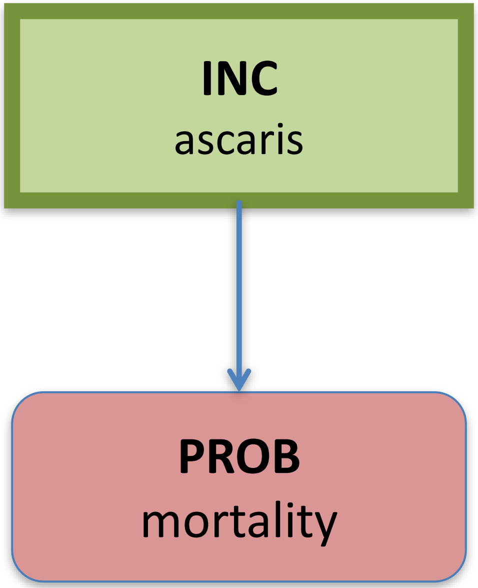

### *Chagas*
### Disease Model

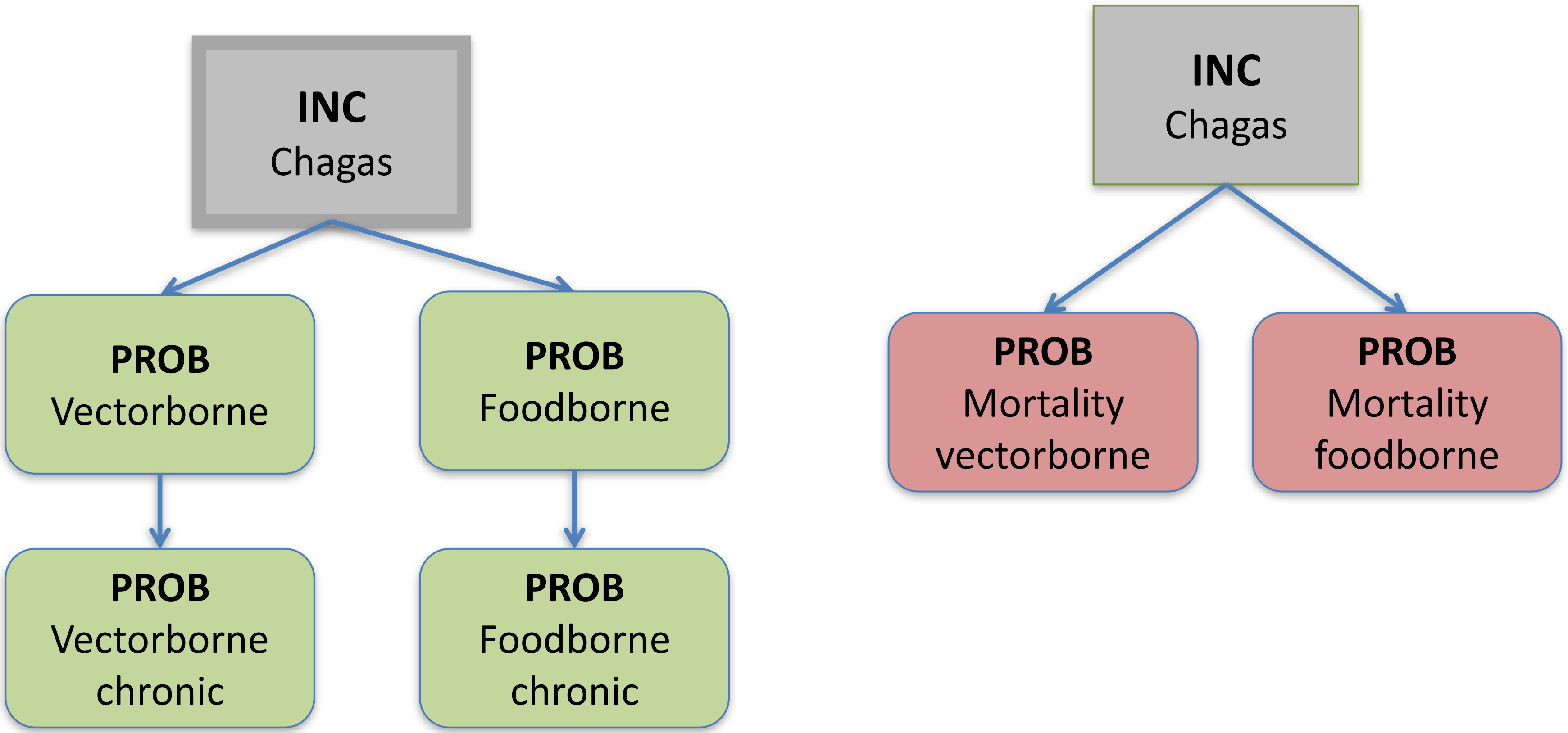

### *Echinococcus granulosus* Disease Model

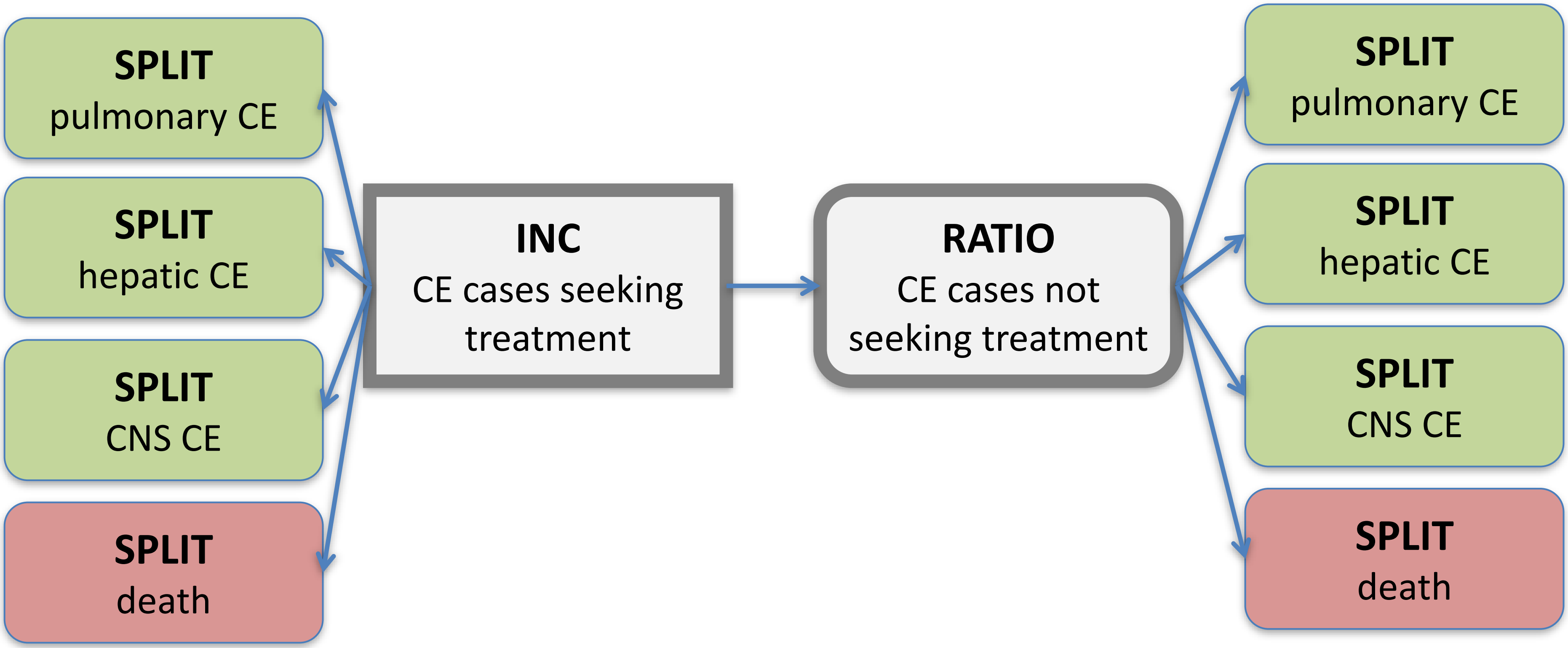

### *Echinococcus multilocularis* Disease Model

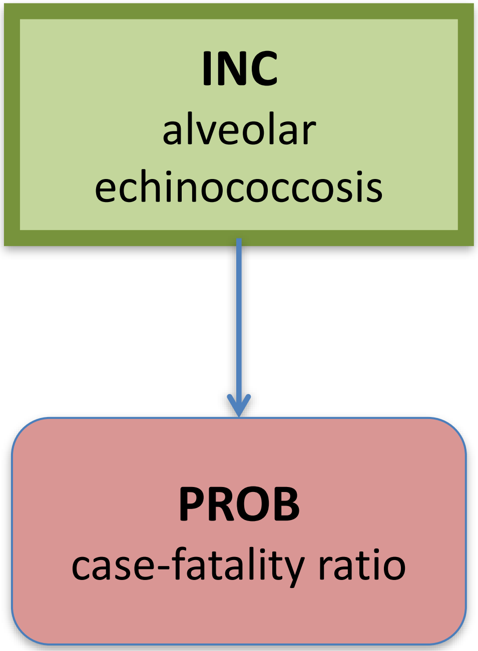

### *Fasciola* spp.
### Disease Model

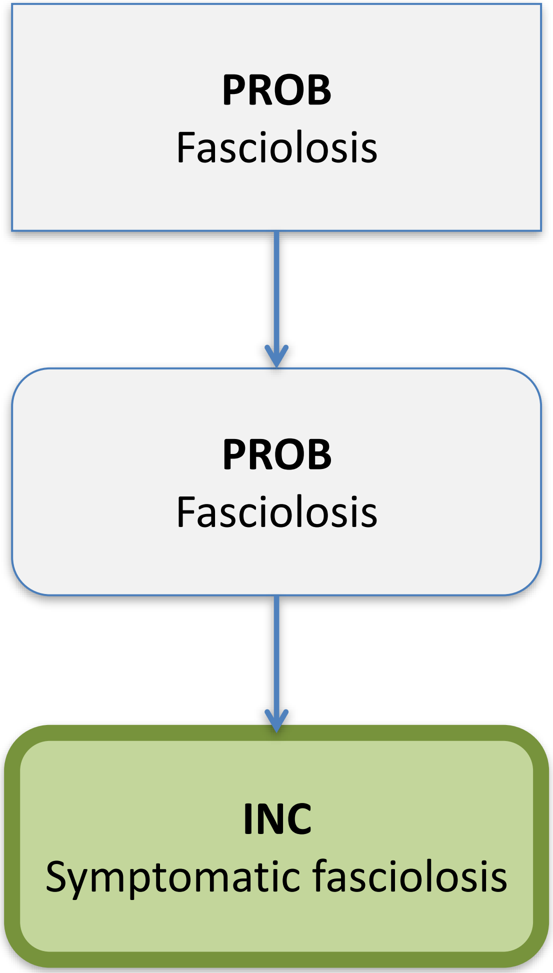

### *Toxoplasma gondii* (acquired) Disease Model

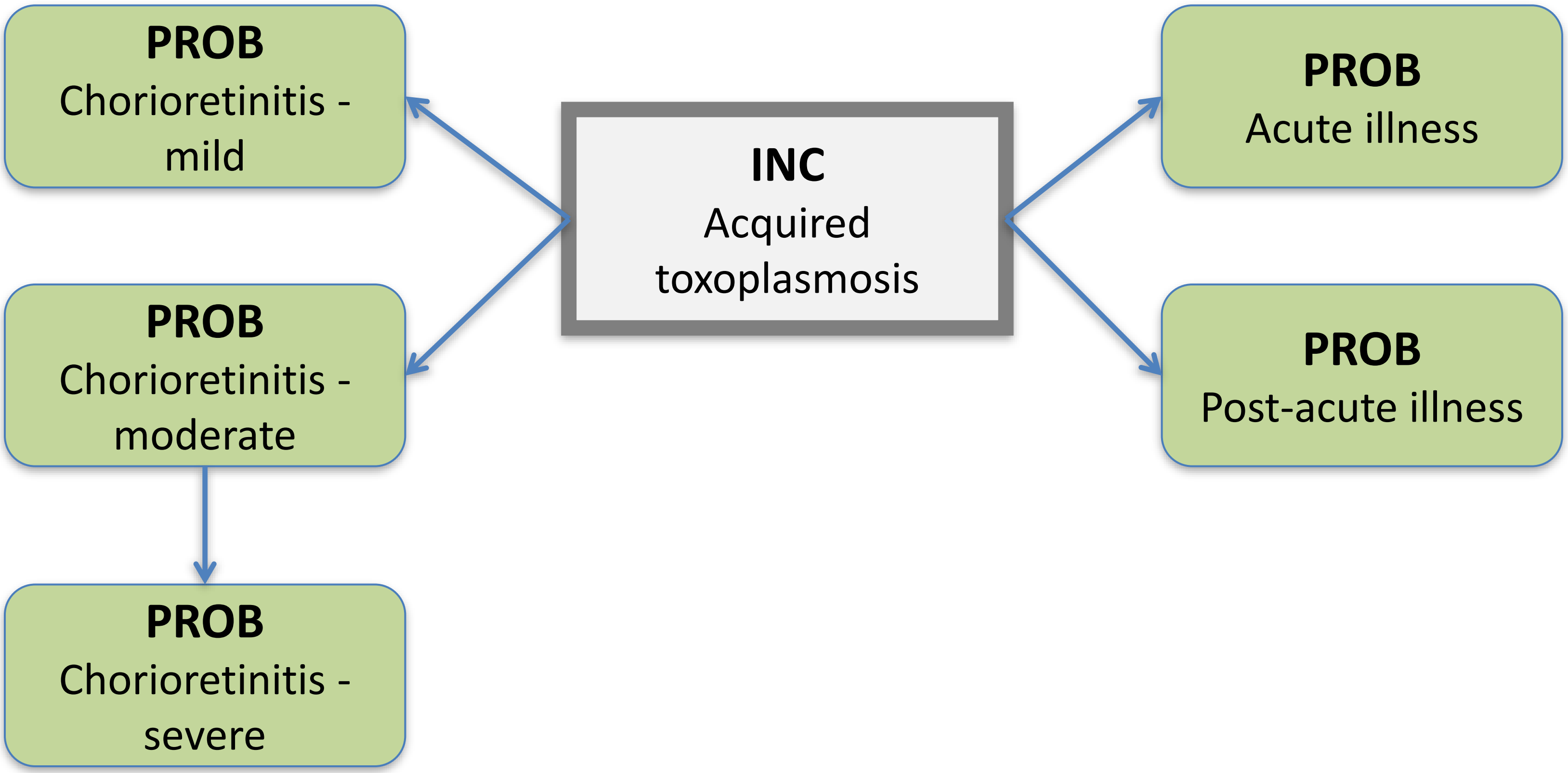

### *Toxoplasma gondii* (congenital) Disease Model

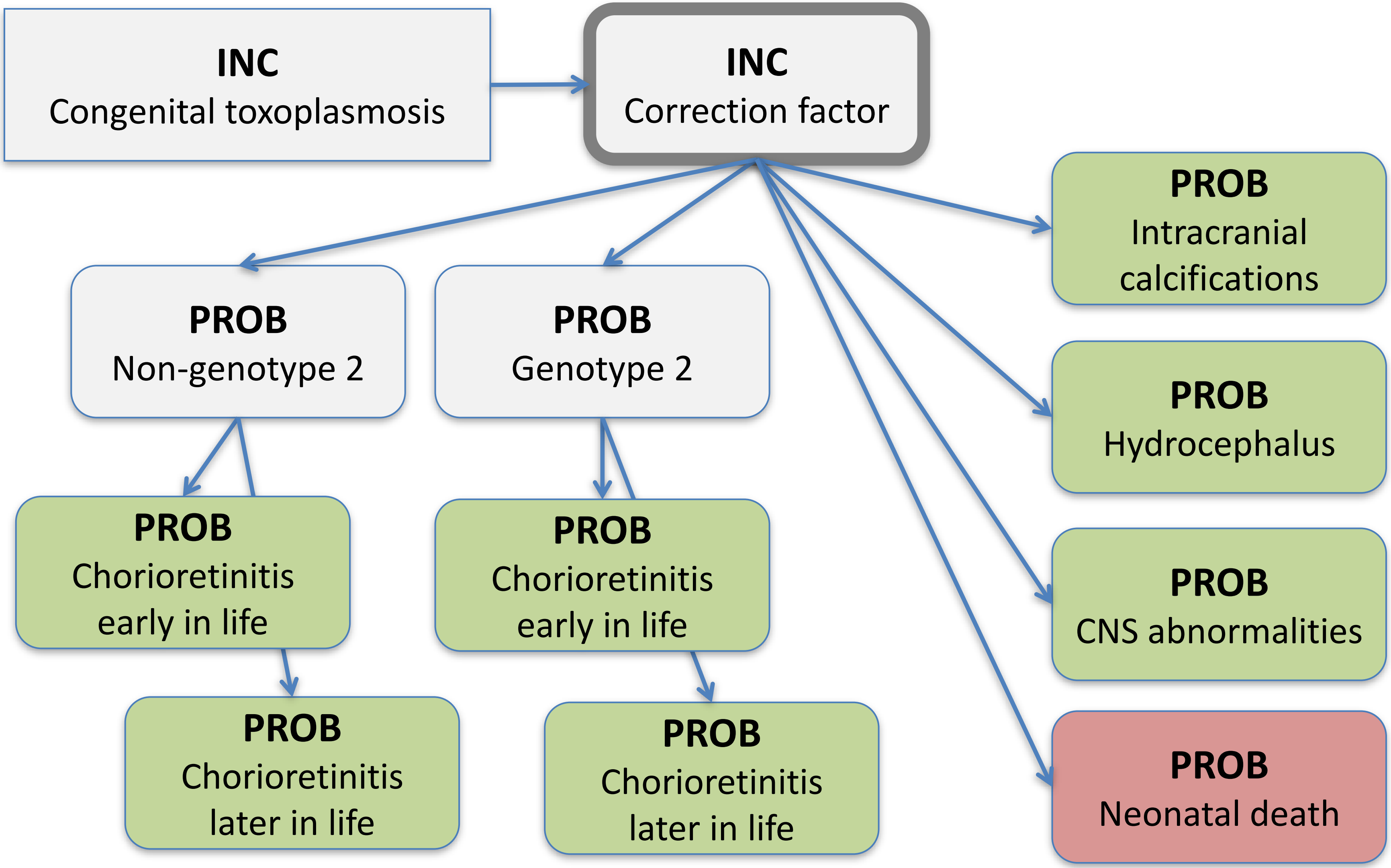

### ***Clonorchis sinensis*** Disease Model

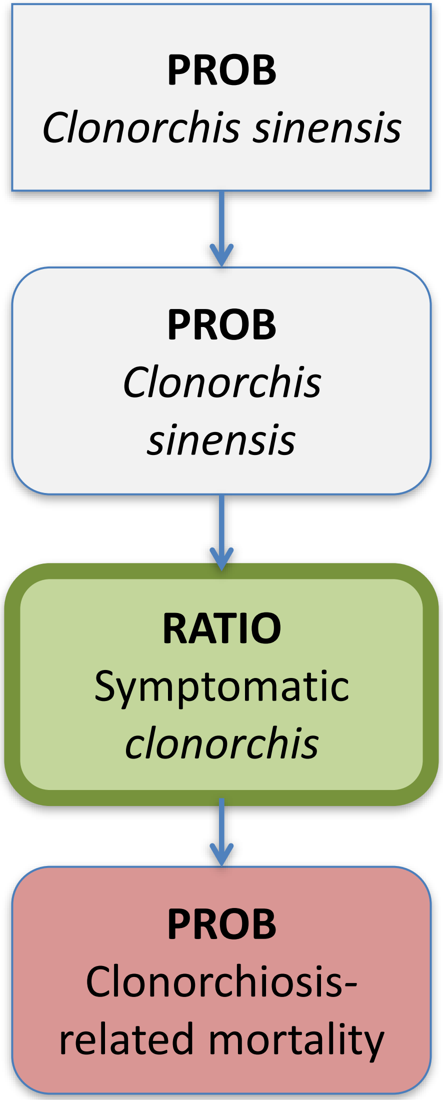

### *Intestinal flukes*
### Disease Model

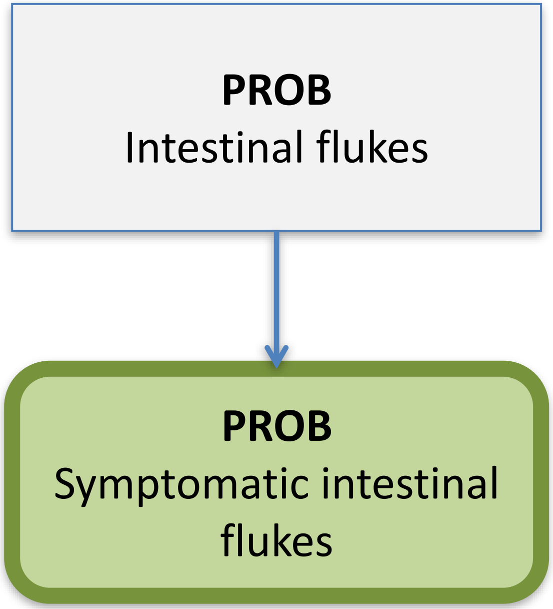

### *Opisthorchis* spp. Disease Model

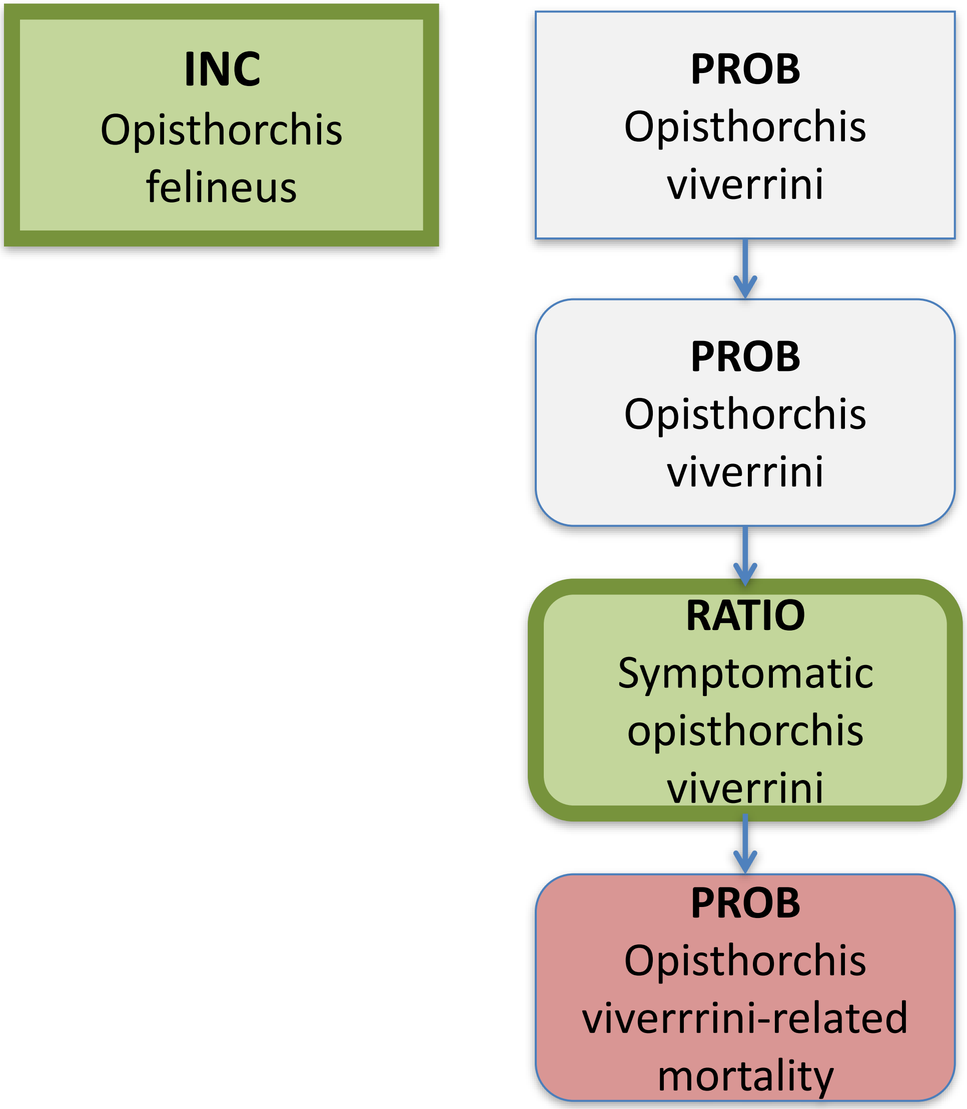

### *Paragonimus*
### Disease Model

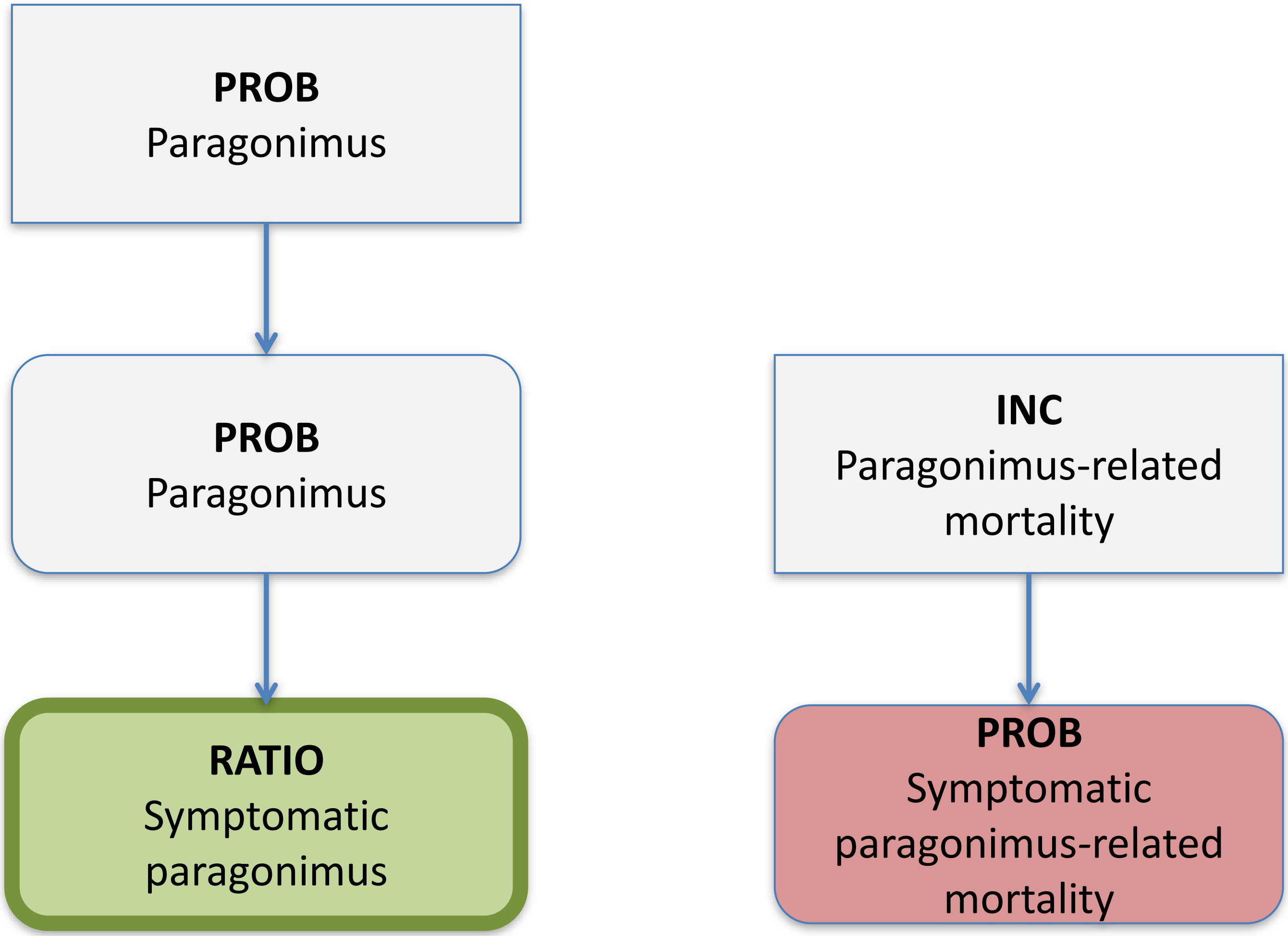

### *Trichinella*
### Disease Model

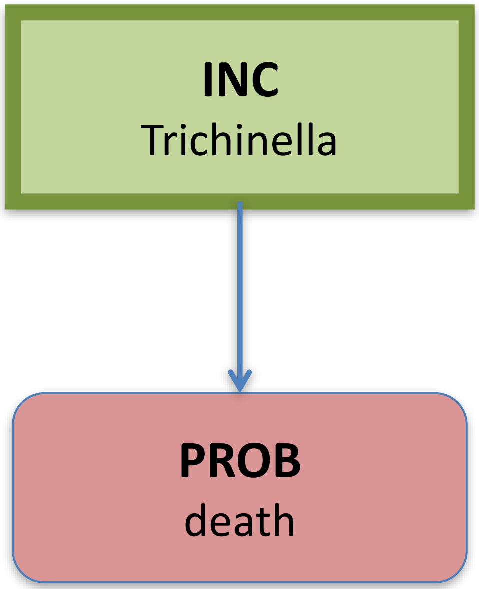

### *Taenia solium*
### Disease Model

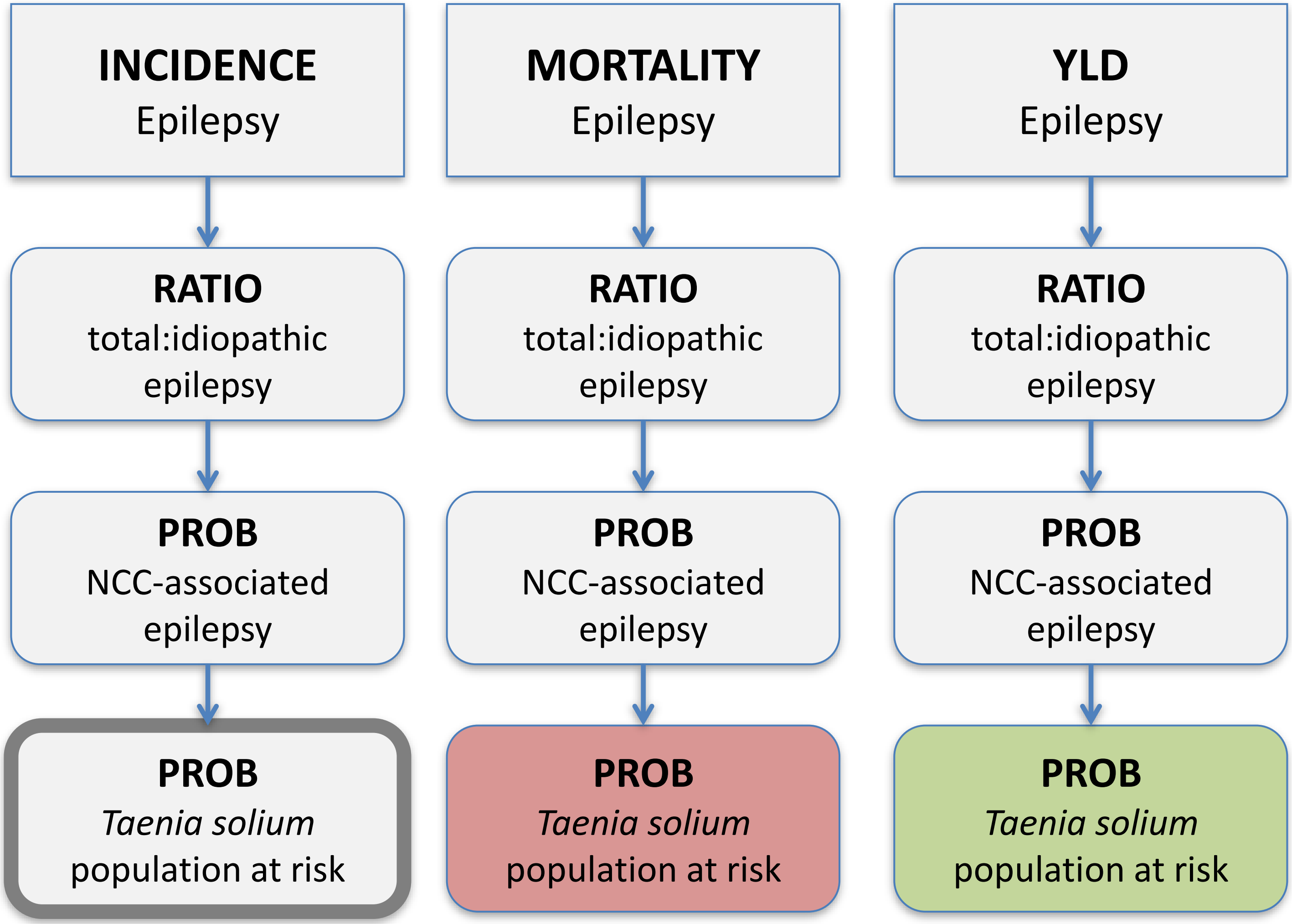
